# Correction for Collider Bias in the Genome-wide Association Study of Diabetes-Related Heart Failure due to Bidirectional Relationship between Heart Failure and Type 2 Diabetes

**DOI:** 10.1101/2023.09.22.23295915

**Authors:** Yan V Sun, Chang Liu, Qin Hui, Jin J Zhou, J Michael Gaziano, Peter WF Wilson, the Million Veteran Program, Jacob Joseph, Lawrence S Phillips

**Author notes:** **Corresponding Author:** Yan V. Sun, PhD Department of Epidemiology, Rollins School of Public Health Emory University 1518 Clifton Rd. NE, Atlanta, Georgia 30322.

## Abstract

**Aims:** Type 2 diabetes (T2D) is a major risk factor for heart failure (HF) across demographic groups. On the other hand, metabolic impairment, including elevated T2D incidence is a hallmark of HF pathophysiology. We investigated the bidirectional relationship between T2D and HF, and identified genetic associations with diabetes-related HF after correction for potential collider bias.

**Methods:** We performed a genome-wide association study (GWAS) of HF to identify genetic instrumental variables (GIVs) for HF, and to enable bidirectional Mendelian Randomization (MR) analysis between T2D and HF. Since genetics and HF can independently influence T2D, collider bias may occur when T2D (i.e., collider) is controlled for by design or analysis. Thus, we conducted GWAS of diabetes-related HF with correction for collider bias.

**Results:** We first identified 61 genomic loci, including 24 novel loci, significantly associated with all-cause HF in 114,275 HF cases and over 1.5 million controls of European ancestry. Combined with the summary statistics of a T2D GWAS, we obtained 59 and 82 GIVs for HF and T2D, respectively. Using a two-sample bidirectional MR approach, we estimated that T2D increased HF risk (OR 1.07, 95% CI 1.04-1.10), while HF also increased T2D risk (OR 1.60, 95% CI 1.36-1.88). Then we performed a GWAS of diabetes-related HF corrected for collider bias due to prevalent HF affecting incidence of T2D. After removing the spurious association of *TCF7L2* locus due to collider bias, we identified two genome-wide significant loci close to *PITX2* (chromosome 4) and *CDKN2B−AS1* (chromosome 9) associated with diabetes-related HF in the Million Veteran Program, and replicated the associations in the UK Biobank study.

**Conclusion:** We identified novel HF-associated loci to enable bidirectional MR study of T2D and HF. Our MR findings support T2D as a HF risk factor and provide strong evidence that HF increases T2D risk. As a result, collider bias leads to spurious genetic associations of diabetes-related HF, which can be effectively corrected to identify true positive loci. Evaluation of collider bias should be a critical component when conducting GWAS of complex disease phenotypes such as diabetes-related cardiovascular complications.

## INTRODUCTION

Heart failure (HF) is a complex life-threatening syndrome that results from structural and functional impairment of ventricular filling or output. HF affects more than 64 million people worldwide ^1^ including 6 million adults in the US^2^. HF prevalence in the US is projected to increase 46% from 2012 to 2030, resulting in over 8 million adults (≥18 years) with HF^3^. In addition to high mortality and morbidity, HF is also associated with high health care costs with an estimated annual expenditure of $70 billion in the US by 2030^2^.

Type 2 Diabetes (T2D) is a complex disease affecting multiple organ systems. The prevalence of T2D has been growing for the past two decades, with age-adjusted prevalence of 9.5% in 1999-2002 and 12% in 2013-2016 among US adults. About 537 million adults live with diabetes around the world, most in low-and middle-income countries^4^. HF is one of the most severe diabetes complications affecting T2D patients’ clinical outcomes and quality of life^5^. Observational studies have consistently demonstrated an increased risk of HF in individuals with DM compared with those without DM, across demographic groups. Even among individuals without T2D, higher levels of fasting glucose and hemoglobin A1c (HbA1c) were associated with increased risk of HF hospitalization^6; 7^. The complex pathogenesis of HF in T2D can include toxic effect of hyperglycemia, diabetic cardiomyopathy, coronary microvascular dysfunction and other comorbid conditions^8^. T2D is associated with a high incidence of both HFpEF and HFrEF^9^, regardless of heterogeneous etiologies, clinical manifestation and outcomes between HF subtypes. Clinical trials have shown that anti-diabetic medications such as sodium-glucose cotransporter-2 (SGLT2) inhibitors can reduce the risk for HF and subtypes^10–12^. Therefore, further understanding of the genetic and molecular mechanisms of diabetes-related HF may lead to therapeutic targets of HF and HF subtypes.

Genome-wide Association Studies (GWAS) are designed to identify genetic loci associated with disease and traits by surveying genome-wide single nucleotide polymorphisms (SNPs). Although no genetic variants have been associated with diabetes-related heart failure, recent GWAS have identified dozens of loci associated with all-cause HF^13–15^ and clinical subtypes including HF with reduced ejection fraction (HFrEF) and HF with preserved ejection fraction (HFpEF)^13^. Genetic and familial studies also estimated the heritability of HF (*h*^2^ ranging from 22% to 34%)^13; 16^ and several diabetes complications including diabetes-related cardiovascular disease (*h*^2^ ∼18%) and diabetes-related stroke (*h*^2^ ∼14%)^17^, which suggested substantial genetic contribution to diabetes-related HF yet to be discovered.

When both exposure (e.g., genetic variants) and outcome (e.g., HF) independently influence a common third variable (e.g., T2D), collider bias can occur when the third variable (i.e., collider) is controlled for by design or analysis. Thus, the bidirectional relationship between T2D and HF can introduce collider bias in the GWAS of diabetes-related HF (both genetics and HF affect T2D). Mendelian Randomization (MR) methods can support such bidirectional relationship using proper instrumental variables of T2D and HF. MR uses genetic variants robustly associated with exposures or risk factors of interest as genetic instrumental variables (GIVs) to estimate the causal and de-confounded relationship between the exposure or risk factor with the disease outcome^18^. Recent genetic studies identified significant but weak MR association between T2D and all-cause HF using two-sample MR approach (OR 1.05-1.08) ^13; 15^, compared to observational studies of T2D and HF. Additionally, the estimated genetic correlation between T2D and all-cause HF of 47.3% ^15^ cannot be fully explained by the moderate MR association between T2D (exposure) and HF (outcome). On the other hand, the hypothesis that HF increases the risk of T2D hasn’t been examined in the MR framework, limited by strong GIVs for HF from large independent samples. In the present study, we identified a large set of GIVs of HF from a GWAS meta-analysis including 114,275 HF cases and 1,506,896 controls of European ancestry. Then we performed the first large scale GWAS of diabetes-related HF, and corrected for collider bias using summary statistics of T2D GWAS to eliminate spurious associations. We also examined and corrected for potential collider bias in T2D-adjusted GWAS of HF.

## SUBJECTS AND METHODS

### Study Samples

The design of the Million Veteran Program (MVP) has been previously described.^19^ Veterans were recruited from over 60 Veterans Health Administration (VA) healthcare systems nationwide since 2011. The MVP links of a large biobank to an extensive electronic health record (EHR) database from 2003 onward that integrates multiple elements such as diagnosis codes, procedure codes, laboratory values, and imaging reports, which permits detailed phenotyping. All MVP participants were genotyped as part of the study design. MVP has received ethical and study protocol approval by the VA Central Institutional Review Board in accordance with the principles outlined in the Declaration of Helsinki. UK Biobank is a prospective study with over 500,000 participants aged 40–69 years recruited in 2006–2010 with extensive phenotypic and genotypic data^20^.

### Phenotypic data

In the MVP, HF patients were identified as those with an International Classification of Diseases (ICD)-9 code of 428.x or ICD-10 code of I50.x and an echocardiogram performed within 6 months of diagnosis (median time period from diagnosis to echocardiography was 3 days, interquartile range 0-32 days)^13^. Based on our previous work, the requirement for echocardiogram improved the specificity of HF diagnosis. The index diagnosis of HF was documented during an outpatient encounter in the majority of cases. We utilized a natural language processing tool to extract LVEF from the VA Text Integration Utilities documents including values measured within and outside the VA^13; 21; 22^. Non-HF controls excluded MVP participants with any recorded HF codes at any time based on their EHR data. Diabetes was defined by both (a) either ≥1 use of the ICD-9 code 250.xx at a primary care provider (PCP) visit, or ≥2 uses of the code in any setting, and (b) an outpatient prescription of a diabetes drug based on use of Veterans Health Administration (VHA) national drug codes^23^.

In the UK Biobank, we defined HF as the presence of self-reported HF, pulmonary edema or cardiomyopathy at any visit; or an ICD-10 or ICD-9 billing code indicative of heart/ventricular failure or a cardiomyopathy of any cause, as described and validated previously, and consistent with that used in recent GWAS of all-cause HF^13; 24^. Similar to the MVP definition, Non-HF controls excluded participants with any self-reported HF or recorded HF codes at any time. Type 2 diabetes (T2D) was defined by the primary and secondary ICD-9 (250 diabetes mellitus, juvenile type excluded) and ICD-10 diagnosis codes (E11 non-insulin-dependent diabetes mellitus, E12 malnutrition-related diabetes mellitus, E13 other specified diabetes mellitus, E14 unspecified diabetes mellitus ^25^), and self-reported T2D at enrollment.

### Genomic data

DNA extracted from participants’ blood was genotyped using a customized Affymetrix Axiom^®^ biobank array. The array was enriched for both common and rare genetic variants of clinical significance in different ethnic backgrounds. Genotype calling, quality-control procedures and genotype imputation were previously described.^26^ We excluded: duplicate samples, samples with more heterozygosity than expected, an excess (>2.5%) of missing genotype calls, or discordance between genetically inferred sex and phenotypic gender.^26^ In addition, one individual from each pair of related individuals (more than second degree relatedness as measured by the KING software^27^) were removed. Prior to imputation, variants that were poorly called (genotype missingness > 5%) or that deviated from their expected allele frequency observed in the 1000 Genomes reference data were excluded. After pre-phasing using EAGLE v2.4^28^, we then imputed to the 1000 Genomes phase 3 version 5 reference panel (1000G) using Minimac4.^29^ Imputed variants with poor imputation quality (r^2^<0.3) were excluded from further analyses.

The MVP participants were assigned to mutually exclusive racial/ethnic groups using HARE (Harmonized Ancestry and Race/Ethnicity), a machine learning algorithm that integrates genetically inferred ancestry (GIA) with self-identified race/ethnicity (SIRE) as previously described.^30^ Briefly, HARE uses GIA to refine SIRE for genetic association studies in three ways: identify individuals whose SIRE are likely inaccurate, reconcile conflicts among multiple SIRE sources, and impute missing racial/ethnic information when the predictive confidence is high. HARE assigned >98% of participants with genotype data to one of four non-overlapping groups: non-Hispanic European (EUR), non-Hispanic African (AFR), Hispanic (HIS), and non-Hispanic Asian Americans (ASN). The present GWAS of diabetes-related HF focused on the MVP EUR group.

To replicate the significant loci associated with diabetes-related HF, we performed a similar genetic association analysis in the UKB participants of European ancestry with available genomic data. Additional sample exclusions were implemented for 3^rd^-degree or closer relatedness (UK Biobank Data Field 22020), sex chromosome aneuploidy, and excess missingness or heterozygosity, as defined by the UK Biobank.

### All-cause HF GWAS meta-analysis

Imputed and directly measured genetic variants from the MVP non-Hispanic European American participants were tested for association assuming an additive genetic model using PLINK2. The GWAS scan included variants with minor allele frequency higher than 1%. Logistic regression of all-cause HF was adjusted for age, sex, and the top ten genotype-derived principal components. We meta-analyzed summary statistics of previously published HF GWAS from the MVP (43,344 cases, 258,943 controls) ^13^, HERMES (47,309 cases, 930,014 controls)^15^ and FinnGen (23,622 cases, 317,939 controls)^31^ studies using the random-effect meta-analysis model implemented in GWAMA^32^. GWAS results were summarized using FUMA, a platform that annotates, prioritizes, visualizes and interprets GWAS results.^33^ Genome-wide significant SNPs (p <5×10^-8^) were grouped into a genomic locus based on either r^2^ > 0.1 or distance between loci of < 500kb using the 1000 Genomes European reference panel. Lead SNPs were defined within each locus if they were independent (r^2^ < 0.1). We considered loci as novel if the sentinel SNP was of genome-wide significance (p <5×10^-8^) and located > 1 Mb from previously reported GWS SNPs associated with HF.

### Genetic instrumental variables (GIVs) for T2D and all-cause HF

We selected independent genetic loci (r^2^ < 0.1) associated with T2D from the large GWAS among participants of European ancestry only or multiple ancestries with predominantly European ancestry participants by 2017^34^. A total of 85 independent T2D-associated SNPs were selected, including SNPs that are genome-wide significant (p < 5×10^-8^) among published GWAS of European ancestry. Among the 85 SNPs, 82 were also present in the all-cause HF GWAS meta-analysis, and thus were used as the GIVs of T2D in the downstream MR analysis. From the all-cause HF GWAS meta-analysis described above, we identified independent genome-wide significant SNPs as the GIVs for HF in the bi-directional MR analysis.

### Two-sample Bidirectional MR

Two-sample MR was conducted to examine possible bidirectional causal associations between T2D and all-cause HF using GIVs from previous GWAS of T2D,^34^ and a large meta-analysis of all-cause HF in the present study. To minimize sample overlap in the two-sample MR design, we used summary statistics of T2D GWAS without UKB and MVP samples, and all-cause HF GWAS from the MVP, HERMES, and FinnGen studies, all from studies of European ancestry. We estimated the MR association between T2D and all-cause HF using three complementary methods: inverse-variance-weighted (IVW), median weighted, and MR-Egger regression, as implemented in the R package *TwoSampleMR*. We reported IVW estimates when the evidence of pleiotropy was not present. MR-Egger regression was used to identify the horizontal pleiotropy indicated by significant intercept of the regression (p-value<0.05). Random-effects model was used to estimate the MR association between exposure and outcome variables for IVW and MR-Egger regression. MR-PRESSO (Mendelian Randomization Pleiotropy RESidual Sum and Outlier) was used to detect and remove outlier GIVs to correct for potential horizontal pleiotropy^35^. As we only evaluate the relationship between T2D and HF, we considered nominal p-value of 0.05 as suggestive evidence for MR association.

Latent Heritable Confounder MR (LHC-MR) is a method designed for analyzing GWAS summary statistics to estimate bidirectional causal effects, while accounting for potential heritable confounder between a pair of traits^36^. LHC-MR can overcome the limitations of traditional MR, including under-exploitation of genome-wide markers, sensitivity to the presence of a heritable confounder, and potential sample overlap^36^. LHC-MR extends the traditional MR model by using a structural equation model incorporating the presence of a latent heritable confounder and estimate its contribution to T2D and all-cause HF separately, while simultaneously estimating the bidirectional causal effect between T2D and all-cause HF. We applied this method to estimate the bidirectional relationship between T2D and all-cause HF using summary statistics from a large T2D GWAS^34^ and the meta-analysis of HF GWAS, both in European ancestry.

### GWAS of diabetes-related HF and T2D-adjusted HF

We conducted GWAS of diabetes-related HF using all-cause HF cases and controls^13^ among 106,321 diabetes patients of European ancestry from the MVP cohort (Table 1). The genetic association of diabetes-related HF was adjusted for age, sex and top 10 PCs. Using the same statistical model, we also performed the GWAS of diabetes-related HF among 26,431 unrelated diabetes patients of European ancestry from the UK Biobank study. We also conducted GWAS of all-cause HF adjusted for T2D status, among 434,089 MVP participants of European ancestry, adjusted for age, sex, T2D status and top 10 PCs.

**Table 1.** Characteristics of the European American participants in the MVP included in the GWAS of diabetic heart failure and T2D-adjusted HF GWAS.

|  | All (n=434,089) |  | T2D (n=106,321) |  | Non-T2D (n=327,768) |  |
| --- | --- | --- | --- | --- | --- | --- |
|  | HF<br>(n= 68,059) | Non-HF<br>Controls<br>(n= 366,030) | HF<br>(n= 31,346) | Non-HF<br>Controls<br>(n= 74,975) | HF<br>(n= 36,713) | Non-HF<br>Controls<br>(n= 291,055) |
| Age, years (SD) | 69.59 (9.628) | 62.49 (14.11) | 68.76 (8.44) | 66.06 (9.80) | 70.29 (10.48) | 61.57 (14.88) |
| Male, n (%) | 65884 (96.80) | 336064 (91.81) | 30509 (97.33) | 71454 (95.30) | 35375 (96.36) | 264610 (90.91) |
| BMI, kg/m<sup>2</sup> (SD) | 31.14 (6.70) | 29.30 (5.54) | 33.33 (6.80) | 31.80 (5.99) | 29.26 (6.01) | 28.66 (5.23) |
| Obesity, (BMI≥30) | 35106 (51.58) | 145631 (39.79) | 20778 (66.29) | 43555 (58.09) | 14328 (39.03) | 102076 (35.07) |
| Atrial fibrillation, n (%) | 22681 (33.33) | 23703 (6.48) | 10232 (32.64) | 6378 (8.51) | 12449 (33.91) | 17325 (5.95) |
| Coronary artery disease, n (%) | 45739 (67.20) | 79154 (21.63) | 23091 (73.66) | 26373 (35.18) | 22648 (61.69) | 52781 (18.13) |
| Chronic kidney disease, n (%) | 23610 (34.69) | 34404 (9.40) | 13615 (43.43) | 13472 (17.97) | 9995 (27.22) | 20932 (7.19) |
| Type 2 Diabetes n (%) | 31346 (46.06) | 74975 (20.48) | 31346 (100) | 74975 (100) | 0 (0) | 0 (0) |
| Hyperlipidemia, n (%) | 59567 (87.52) | 244010 (66.66) | 29724 (94.83) | 67780 (90.40) | 29843 (81.29) | 176230 (60.55) |
| Hypertension, n (%) | 62596 (91.97) | 228065 (62.31) | 30573 (97.53) | 67346 (89.82) | 32023 (87.23) | 160719 (55.22) |

T2D: Type-2 Diabetes; HF: Heart Failure; SD: Standard Deviation; n: number; BMI: Body Mass Index.
All p-values comparing HF vs. non-HF controls were $<2.2 \times 10^{-16}$ except for Type 2 Diabetes in T2D-stratified subgroups (not available).

### Correction for collider bias using Slope-Hunter

The Slope-Hunter method was developed for correcting collider bias in conditional GWAS using genetic effects of the collider (i.e., T2D) and the outcome variable (i.e., HF) ^37^. It uses model-based clustering to estimate the adjustment factor for GWAS results. We obtained GWAS summary statistics for diabetes-related HF and T2D from the MVP study, and considered 7,700,660 variants (MAF>0.01) present in both datasets. An independent set of SNPs was obtained after performing LD-pruning using PLINK2 software (R^2^ threshold of 0.1 within 250-SNP windows). The LD-pruning was estimated using the European ancestry population of the 1000 Genomes reference panel. The results obtained from the Slope-Hunter method were compared with results from the naive GWAS of T2D-related HF, and T2D-adjusted HF.

## RESULTS

The present study consists of a large meta-analysis of all-cause HF in the European ancestry to enable the bidirectional MR study of T2D and HF, followed by a GWAS of diabetes-related HF with collider bias correction (Figure 1). The primary study population consisted of 106,321 MVP participants with T2D diagnosis, out of 434,089 with European ancestry, predominantly male. In the GWAS of diabetes-related HF, we include 31,346 HF cases with comorbid T2D, and 74,975 non-HF diabetes controls (Table 1). HF patients were older, and had higher prevalence of obesity, atrial fibrillation, coronary artery disease, chronic kidney disease, hyperlipidemia and hypertension with or without T2D (Table 1). The prevalence of all-cause HF was higher among T2D patients (29.5%) than that among non-diabetes participants (11.2%). In the UK Biobank, we included 26,431 T2D patients with European ancestry. Among them, 3,506 developed HF using clinical diagnosis codes (Supplementary Table 1). Similarly, HF patients had significant (p<0.001) older age, higher prevalence of cardiometabolic risk factors and comorbidities than the control populations without HF.

**Figure 1.**
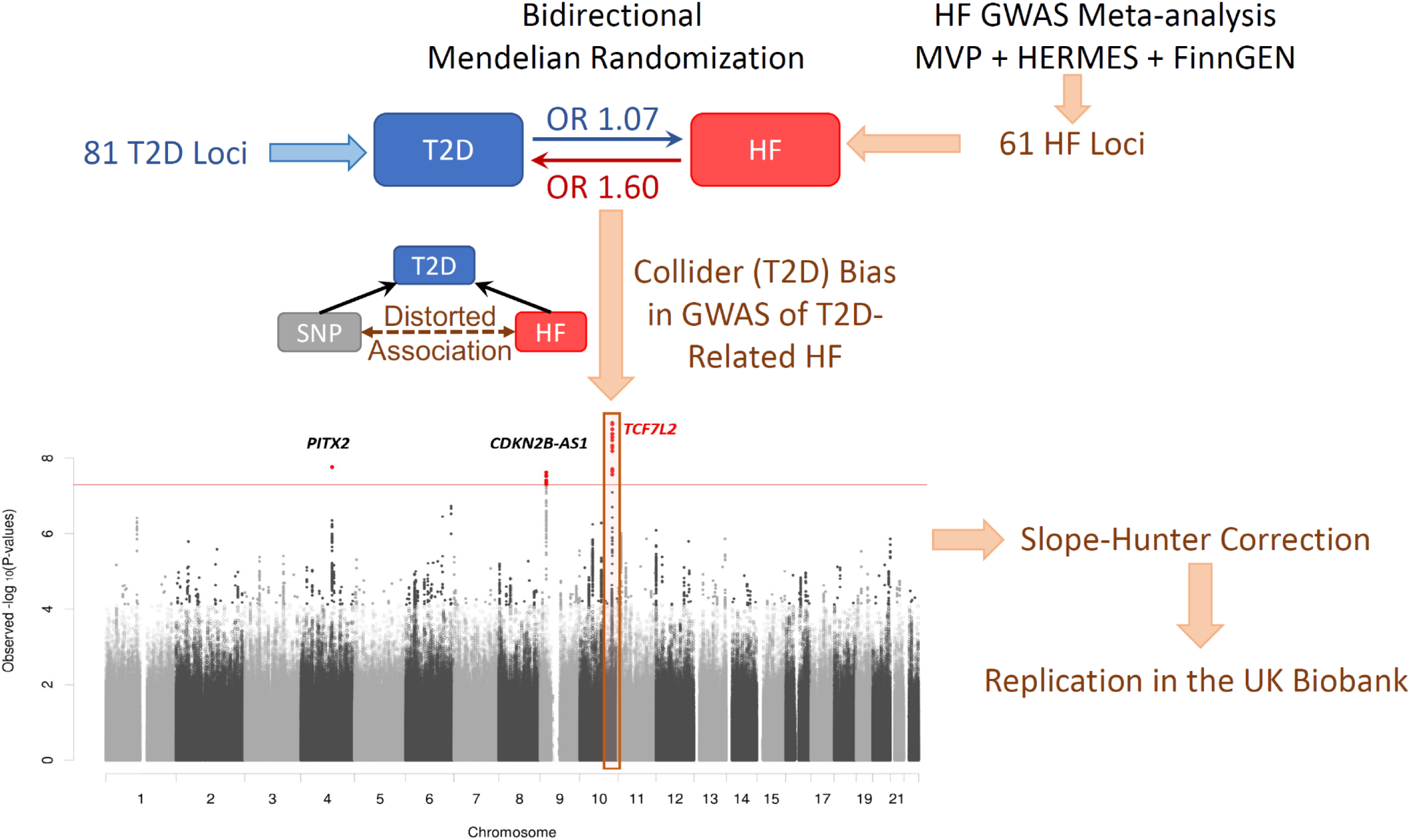
Overview of study design.

### Genome-wide meta-analysis of all-cause HF

A total of 10,835,443 SNPs with MAF > 1% in any one of the three studies (i.e., MVP, HERMES and FinnGen) were included in the meta-analysis of all-cause HF among European ancestry. We identified a total of 61 independent genome-wide significant loci (Supplementary Table 2) associated with all-cause HF, including 24 novel loci (Table 2) comparing to previous reported HF GWAS^13–15^. Overlapping with a T2D GWAS^34^, 59 out of 61 HF-associated SNPs also had summary statistics, and were used as the GIVs for all-cause HF in the two-sample MR analysis (Supplementary Table 3).

**Table 2.**
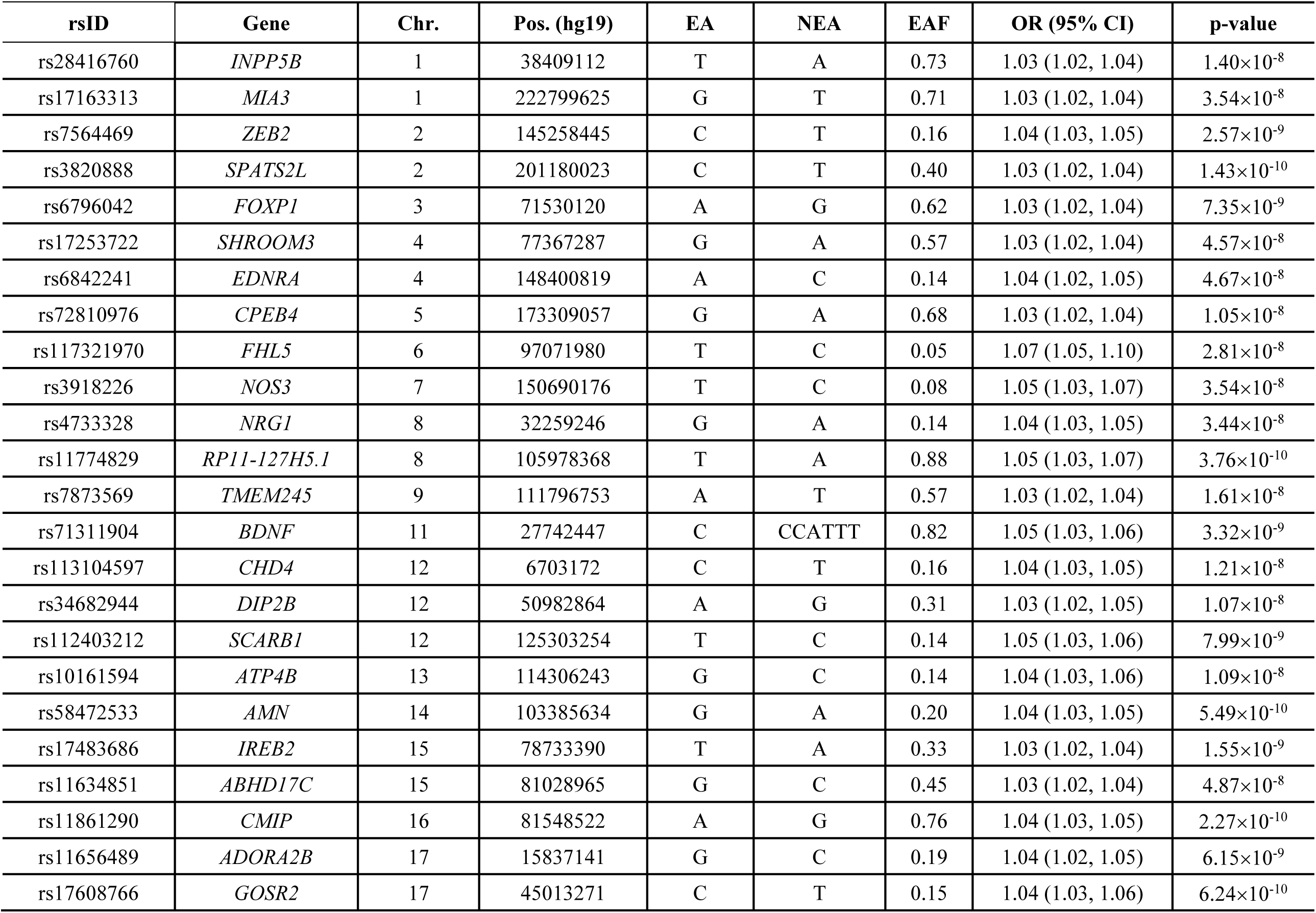
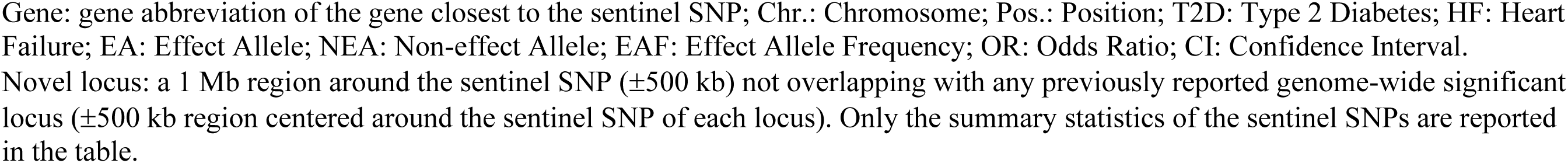
Twenty-four novel genome-wide significant loci associated with all-cause heart failure.

### Bidirectional MR analysis between T2D and all-cause HF

A total of 82 GIVs for T2D (Supplementary Table 4) and 59 for HF had summary statistics in both T2D and HF GWAS. IVW-MR method showed significant MR association in both directions (Figure 2, Supplementary Table 5), suggesting potential causal effect of T2D on HF (OR 1.07, 95% CI 1.04-1.10, p=7.02×10^-7^)), as well as potential causal effect of HF on T2D (OR 1.60, 95% CI 1.36-1.88, p=1.55×10^-8^). The MR-Egger method didn’t support a significant intercept which indicates limited pleiotropy. Therefore, the IVW estimates were largely accurate. Only the HF effect on T2D showed significant positive association in MR-Egger analysis. After removing 5 (rs10965223, rs635634, rs7903146, rs1061810, rs1558902) and 2 (rs600038, rs11642015) outliers for T2D and HF, respectively, the MR-PRESSO analysis showed similar significant MR associations between T2D and HF in both directions using IVM and MR-Egger methods (Figure 2, Supplementary Table 5).

**Figure 2.**
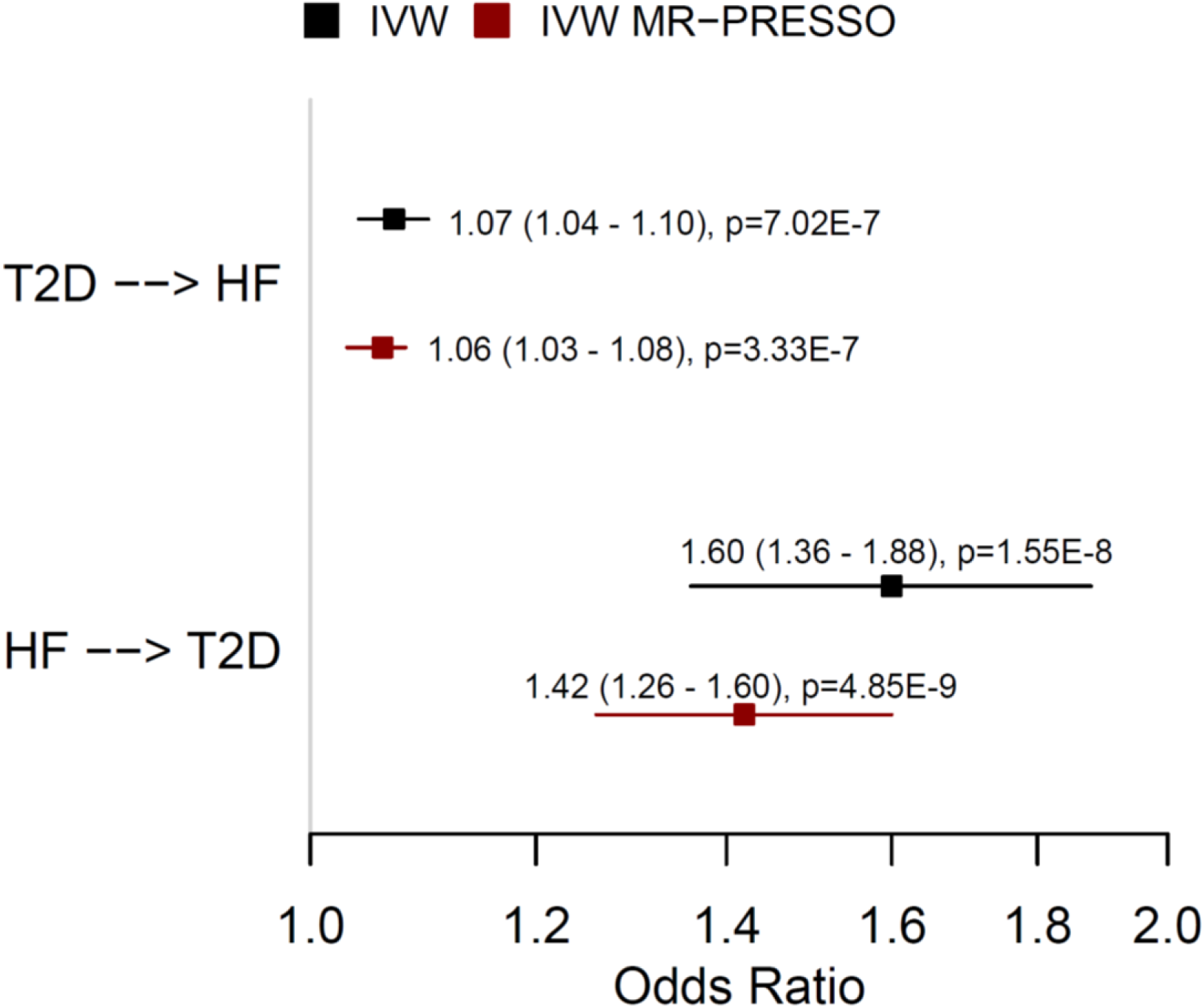
Forest plot of bidirectional Mendelian Randomization between T2D and all-cause HF. 95% confidence interval of odds ratio is included in the parentheses. IVW: inverse-variance weighted; MR: Mendelian Randomization; p: p-value.

Using LHC-MR method, we also identified bi-directional relationship between all-cause HF and T2D. Similar to two-sample MR results, T2D is associated with higher risk for HF with moderate effect size (OR 1.09, 95% CI 1.06-1.13, p-value 3.95×10^-7^). Meanwhile, HF is associated with higher risk for T2D with much larger effect size (OR 1.95, 95% CI 1.55-2.44, p-value 6.82×10^-9^).

### GWAS of Diabetes-Related HF with Slope-Hunter Correction

In the GWAS of diabetes-related HF among 106,321 diabetic patients (31,346 all-cause HF cases, 29.5%), we identified nine suggestively significant (p-value<10^-6^) loci including three GWS (p-value<5×10^-8^) loci associated with diabetes-related HF (Table 3, Figure 3). The inflation factor of the GWAS is 1.04. One diabetes-related HF-associated locus located on chromosome 10 (*TCF7L2*) is strongly associated with T2D but not associated with all-cause HF in the meta-analysis (p-value of 0.70), which can be affected by collider bias. After Slope-Hunter correction, the *TCF7L2* locus was no longer associated with diabetes-related HF (OR 1.02, 95% CI 0.99-1.05, p-value 0.15). Meanwhile, the other two loci on chromosome 4 (sentinel SNP rs17513625 close to *PITX2*, OR 1.25, 95% CI 1.16-1.35, p-value 9.98×10^-9^) and 9 (sentinel SNP rs4977575 close to *CDKN2B−AS1*, OR 1.06, 95% CI 1.04-1.08, p-value 2.91×10^-9^) remained GWS after Slope-Hunter correction for collider bias. Interestingly, the genetic association of chromosome 4 locus with HF was much weaker among 327,768 MVP participants without T2D (OR 1.07, 95% CI 1.00-1.14, p-value 0.039), indicating the modification effect of T2D (interaction p-value 0.016). We pursued replication of two GWS loci on chromosome 4 and 9 using the UK Biobank study participants with European ancestry (Supplementary Table 1). After applying the Slope-Hunter correction to the GWAS of diabetes-related HF adjusted for age, sex and top ten PCs, consistent associations were identified for rs17513625 (*PITX2* locus, OR 1.19, 95% CI 1.02-1.40, p-value 0.027), and rs4977575 (*CDKN2B−AS1* locus, OR 1.08, 1.03-1.14, p-value 0.0034), respectively.

**Figure 3.**
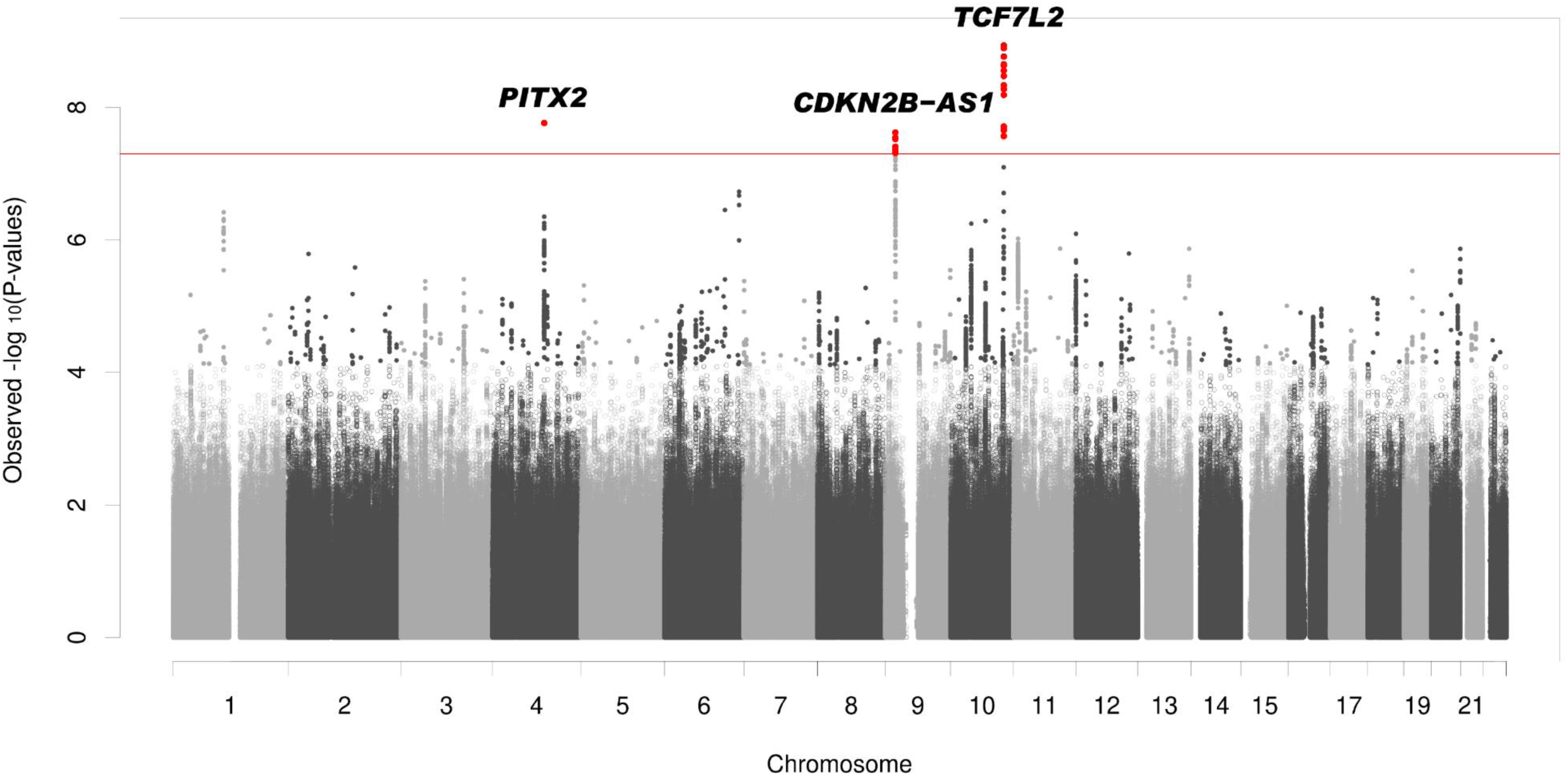
Manhattan plot of diabetes-related HF GWAS without correction for collider bias. Red horizontal line indicates genome-wide significance threshold of nominal p-value of 5×10^-8^.

**Table 3.** Genomic loci associated with diabetes-related heart failure (genomic loci with p<10^-6^) with correction for collider bias using Slope-Hunter.

| rsID | Gene | Chr | Position | EA | NEA | EAF | diabetes-related HF GWAS |  | T2D GWAS |  | diabetes-related HF GWAS after Slope-Hunter Correction |  | HF GWAS Meta-analysis |  |
| --- | --- | --- | --- | --- | --- | --- | --- | --- | --- | --- | --- | --- | --- | --- |
|  |  |  |  |  |  |  | OR (95% CI) | P | OR (95% CI) | P | OR (95% CI) | P | OR (95% CI) | P |
| rs602633 | *PSRC1* * | 1 | 109821511 | G | T | 0.78 | 1.06 (1.04, 1.09) | $3.83 \times 10^{-7}$ | 1 (0.99, 1.02) | 0.518 | 1.06 (1.04, 1.09) | $3.11 \times 10^{-7}$ | 1.05 (1.04, 1.06) | $6.75 \times 10^{-17}$ |
| **rs17513625** | ***PITX2*\*** | **4** | **111848270** | **A** | **G** | **0.02** | **1.24 (1.15, 1.34)** | **$1.72 \times 10^{-8}$** | **1.03 (0.98, 1.08)** | **0.280** | **1.25 (1.16, 1.35)** | **$9.98 \times 10^{-9}$** | **1.11 (1.08, 1.14)** | **$6.63 \times 10^{-14}$** |
| rs55730499 | *LPA* * | 6 | 161005610 | T | C | 0.07 | 1.11 (1.07, 1.15) | $1.87 \times 10^{-7}$ | 1 (0.97, 1.02) | 0.767 | 1.11 (1.06, 1.15) | $3.02 \times 10^{-7}$ | 1.1 (1.08, 1.12) | $1.79 \times 10^{-23}$ |
| **rs4977575** | ***CDKN2B-AS1*\*** | **9** | **22124744** | **G** | **C** | **0.5** | **1.06 (1.04, 1.08)** | **$2.40 \times 10^{-8}$** | **1.02 (1.01, 1.03)** | **$1.74 \times 10^{-3}$** | **1.06 (1.04, 1.08)** | **$2.91 \times 10^{-9}$** | **1.06 (1.05, 1.07)** | **$1.37 \times 10^{-31}$** |
| rs1837530484 | *LINC02881* | 10 | 44738619 | CA | C | 0.91 | 1.1 (1.06, 1.14) | $5.68 \times 10^{-7}$ | 1 (0.97, 1.02) | 0.681 | 1.09 (1.06, 1.14) | $9.37 \times 10^{-7}$ | 1.02 (1, 1.05) | 0.0306 |
| rs201426892 | *AGAP5* | 10 | 75439094 | G | A | 0.99 | 1.78 (1.42, 2.24) | $5.17 \times 10^{-7}$ | 1.07 (0.94, 1.21) | 0.287 | 1.81 (1.44, 2.27) | $3.23 \times 10^{-7}$ | NA | NA |
| ***rs11196211*** | ***TCF7L2*** | **10** | **114817009** | **A** | **C** | **0.69** | **1.07 (1.05, 1.1)** | **$1.16 \times 10^{-9}$** | **0.8 (0.79, 0.81)** | **$2.50 \times 10^{-235}$** | **1.02 (0.99, 1.05)** | **0.154** | **1 (0.99, 1.01)** | **0.569** |
| rs4403799 | *AMPD3* | 11 | 10330455 | G | A | 0.11 | 1.08 (1.05, 1.12) | $9.58 \times 10^{-7}$ | 0.99 (0.97, 1.01) | 0.497 | 1.08 (1.05, 1.12) | $2.05 \times 10^{-6}$ | 1.03 (1.02, 1.05) | $5.57 \times 10^{-06}$ |
| rs797765 | *SLC6A13* | 12 | 372438 | G | A | 0.78 | 1.06 (1.04, 1.09) | $8.12 \times 10^{-7}$ | 0.98 (0.96, 0.99) | $2.76 \times 10^{-3}$ | 1.06 (1.03, 1.08) | $7.61 \times 10^{-6}$ | 1.02 (1.01, 1.03) | 0.0016 |

Chr: Chromosome; Position: human genome build hg19; EA: Effect Allele; NEA: Non-Effect Allele; EAF: Effect Allele Frequency; HF: Heart Failure; GWAS: Genome-wide Association Study; T2D: Type 2 Diabetes; OR: Odds Ratio; CI: Confidence Interval. Bold font: genome-wide significant association ( $p$ -value $< 5 \times 10^{-8}$ ) before and after Slope-Hunter correction; Bold italic font: genome-wide significant association ( $p$ -value $< 5 \times 10^{-8}$ ) with diabetes-related HF but not significant ( $p$ -value $> 0.05$ ) after Slope-Hunter correction. \*: GWS loci associated with all-cause HF in the meta-analysis of the European ancestry.

## DISCUSSION

The present study aimed to elucidate the relationship between T2D and HF, and identify the genetic loci of T2D-related HF. Using GWS loci from a large meta-analysis of all-cause HF, we conducted a bidirectional MR analysis to investigate the relationship between T2D and HF. The estimates from the two-sample MR strongly supported that not only T2D is a risk factor of HF, but also HF increases the risk for T2D. As a result, a T2D-stratified or a T2D-adjusted HF GWAS may identify spurious genetics associations due to collider bias (both HF and genetic factors can affect T2D). We adopted a recently developed method, Slope-Hunter, to correct for such collider bias in the GWAS of T2D-related GWAS among over 100,000 T2D patients from the MVP. After removing the T2D-associated *TCF7L2* locus by Slope-Hunter correction, we identified two GWS loci associated with T2D-related HF, located on chromosome 4 (*PITX2*) and chromosome 9 (*CDKN2B−AS1*). Although both loci have been associated with all-cause HF ^13;14^, the effect size of the SNP (*PITX2*) was larger among T2D patients than that among non-T2D participants. In addition, the sentinel SNP rs17513625 is weakly correlated with the established atrial fibrillation-associated *PITX2* locus (LD R^2^ of 0.115 with rs17042175 in the European ancestry). By definition of collider bias, we anticipated that the collider bias could also affect the T2D-adjusted GWAS of all-cause HF. Without Slope-Hunter correction, we identified GWS 22 loci associated with all-cause HF among non-Hispanic White participants (Supplementary Table 6, Supplementary Figure 2). Two loci located on chromosome 1 (*C1orf185*) and chromosome 10 (*TCF7L2*) were not GWS after Slope-Hunter correction. Both loci were significantly associated with T2D (Supplementary Table 6).

Observational studies consistently demonstrated that diabetes increases the risk for HF. On the other hand, HF induces metabolic impairment, which leads to higher incidence of T2D among HF patients than in comparable general populations. Significant MR associations from the present study supported the bidirectional causal relationship between T2D and all-cause HF. Regardless of the directionality of the effects, adults with both diabetes and heart failure can have 8.8 folds higher mortality rate than those without heart failure (32.7 vs. 3.7 per 1000 person-years)^38^. Thus, managing T2D and hyperglycemia can be effective to prevent HF, to mitigate T2D progression, and eventually reduce mortality among HF patients. *SGLT2* inhibitors are a new class of antidiabetic medications which reduces hyperglycemia through inhibition of glucose reabsorption in the renal proximal tubules. They significantly reduced the risk of HF-related hospitalization and cardiovascular death^10–12^. SGLT2 inhibitors are recommended for patients HF irrespective of diabetes status ^39^.

Both T2D and HF are complex clinical conditions involving numerous risk factors and pathways. Recent studies identified subtypes of T2D using risk factor and biomarker data, that presented differential clinical outcomes^40; 41^. Analyses of T2D-associated loci also revealed genetic clusters linking with pathophysiological pathways underlying T2D^42^, supporting the heterogeneity of T2D mechanism. One the other hand, the heterogeneity of HF has been well documented, even among the major clinical subtypes. Based on the measurement of LVEF, recently guideline categorized HF into HFrEF, HFpEF, HF with mildly reduced EF (HFmrEF) and improved EF (HFimpHF), with HFrEF and HFpEF as the dominant forms^39^. Not surprisingly, HFrEF and HFpEF have distinguishable risk profiles, different response to treatments, and contrasting clinical prognosis. Even within HFpEF subtype, the evidence of heterogeneous subtypes has emerged to support precision treatment and prognosis ^43^, which holds the promise for mitigating the growing burden of HFpEF in the aging population. A recent large GWAS of HFrEF and HFpEF also highlighted the different genetic architecture between two HF clinical subtypes, and supported the phenotypic heterogeneity of HFpEF^13^. However, the limited number of HF subtype GWAS and identified loci, the power of the bidirectional MR between T2D and HF subtypes, and the GWAS are suboptimal. Particularly, only one HFpEF-associated loci close to *FTO* gene has been reported. Since the *FTO* locus is highly pleiotropic, it cannot be used as a GIV of HFpEF in the MR analysis. Future GWAS of HF subtypes would provide more GIVs to robustly estimate the relationship between T2D and HF subtypes, and to accurately identify genetic loci of HF with comorbid conditions with correction of potential collider bias.

## CONCLUSION

Global trend of growing T2D and HF requires improved intervention and prevention strategies for diabetes-related HF, a syndrome with high morbidity and mortality. Exploring the genetic architecture of diabetes-related HF would greatly help understand the mechanism and pathophysiology of the condition as shown in recent GWAS of human diseases. However, the complexity of genetic factors underlying T2D and HF, as well as the relationship between them, created a unique challenge in the identification of true genetic associations with diabetes-related HF. We have demonstrated the evidence supporting the bidirectional relationship between T2D and HF, addressed the impact of collider bias on the GWAS of diabetes-related HF, identified and replicated two genetic loci in the MVP and UK Biobank, two large biobank studies. The study design and analytical workflow can be extended to other studies of diabetic complications, particularly outcomes related to HF and HF subtypes. In light of growing precision medicine studies focusing on certainly disease subgroups or patients with specific comorbid conditions, this case study presented the key considerations of epidemiologic, genetic and biostatistical evidence and methods for such complex disease research in target populations.

## Data Availability

Due to US Department of Veterans Affairs (VA) regulations and our ethics agreements, the analytic datasets used for this study are not permitted to leave the Million Veteran Program (MVP) research environment and VA firewall. This limitation is consistent with other MVP studies based on VA data. However, the MVP data are made available to researchers with an approved VA and MVP study protocol. The full summary level association data genome-wide association analyses in the MVP and the meta-analysis from this report will be available through dbGaP (accession number phs001672). The only restriction is that use of the data is limited to health/medical/biomedical purposes, and does not include the study of population origins or ancestry. Use of the data does include methods development research (e.g., development and testing of software or algorithms) and requestors agree to make the results of studies using the data available to the larger scientific community. We used publicly available data from GTEx (https://gtexportal.org/home/), and FinnGEN (https://www.finngen.fi/en/access_results).
We utilized publicly available software for all analyses, and software used in this study is described in the Methods section.

## Acknowledgement

We are grateful to all the MVP investigators; a list of MVP investigators can be found in Supplementary Materials. This research has been conducted using the UK Biobank Resource under Application Number “34031”.

## Author Contributions

Y.V.S conceptualized the research idea, supervised the study, and wrote the original draft. C.L and H.Q. analyzed data and contributed to manuscript writing. J.J.Z., J.M.G, P.W.F.W., J.J., and L.S.P. contributed to manuscript review and editing. All authors contributed to discussions about the results and provided feedback on the manuscript.

## Data and Code Availability

Due to US Department of Veterans Affairs (VA) regulations and our ethics agreements, the analytic datasets used for this study are not permitted to leave the Million Veteran Program (MVP) research environment and VA firewall. This limitation is consistent with other MVP studies based on VA data. However, the MVP data are made available to researchers with an approved VA and MVP study protocol. The full summary level association data genome-wide association analyses in the MVP and the meta-analysis from this report will be available through dbGaP (accession number phs001672). The only restriction is that use of the data is limited to health/medical/biomedical purposes, and does not include the study of population origins or ancestry. Use of the data does include methods development research (e.g., development and testing of software or algorithms) and requestors agree to make the results of studies using the data available to the larger scientific community. We used publicly available data from GTEx (https://gtexportal.org/home/), and FinnGEN (https://www.finngen.fi/en/access_results). We utilized publicly available software for all analyses, and software used in this study is described in the Methods section.

## REFERENCES

1. Savarese, G., Becher, P.M., Lund, L.H., Seferovic, P., Rosano, G.M.C., and Coats, A.J.S. (2023). Global burden of heart failure: a comprehensive and updated review of epidemiology. Cardiovasc Res 118, 3272–3287.

2. Virani, S.S., Alonso, A., Aparicio, H.J., Benjamin, E.J., Bittencourt, M.S., Callaway, C.W., Carson, A.P., Chamberlain, A.M., Cheng, S., Delling, F.N., et al. (2021). Heart Disease and Stroke Statistics-2021 Update: A Report From the American Heart Association. Circulation 143, e254-e743.

3. Heidenreich, P.A., Albert, N.M., Allen, L.A., Bluemke, D.A., Butler, J., Fonarow, G.C., Ikonomidis, J.S., Khavjou, O., Konstam, M.A., Maddox, T.M., et al. (2013). Forecasting the impact of heart failure in the United States: a policy statement from the American Heart Association. Circ Heart Fail 6, 606–619.

4. IDF Diabetes Atlas. https://diabetesatlas.org/.

5. Dunlay, S.M., Givertz, M.M., Aguilar, D., Allen, L.A., Chan, M., Desai, A.S., Deswal, A., Dickson, V.V., Kosiborod, M.N., Lekavich, C.L., et al. (2019). Type 2 Diabetes Mellitus and Heart Failure: A Scientific Statement From the American Heart Association and the Heart Failure Society of America: This statement does not represent an update of the 2017 ACC/AHA/HFSA heart failure guideline update. Circulation 140, e294–e324.

6. Matsushita, K., Blecker, S., Pazin-Filho, A., Bertoni, A., Chang, P.P., Coresh, J., and Selvin, E. (2010). The association of hemoglobin a1c with incident heart failure among people without diabetes: the atherosclerosis risk in communities study. Diabetes 59, 2020–2026.

7. Held, C., Gerstein, H.C., Yusuf, S., Zhao, F., Hilbrich, L., Anderson, C., Sleight, P., Teo, K., and Investigators, O.T. (2007). Glucose levels predict hospitalization for congestive heart failure in patients at high cardiovascular risk. Circulation 115, 1371–1375.

8. Triposkiadis, F., Xanthopoulos, A., Bargiota, A., Kitai, T., Katsiki, N., Farmakis, D., Skoularigis, J., Starling, R.C., and Iliodromitis, E. (2021). Diabetes Mellitus and Heart Failure. J Clin Med 10.

9. Kodama, S., Fujihara, K., Horikawa, C., Sato, T., Iwanaga, M., Yamada, T., Kato, K., Watanabe, K., Shimano, H., Izumi, T., et al. (2020). Diabetes mellitus and risk of new-onset and recurrent heart failure: a systematic review and meta-analysis. ESC Heart Fail 7, 2146–2174.

10. Zinman, B., Wanner, C., Lachin, J.M., Fitchett, D., Bluhmki, E., Hantel, S., Mattheus, M., Devins, T., Johansen, O.E., Woerle, H.J., et al. (2015). Empagliflozin, Cardiovascular Outcomes, and Mortality in Type 2 Diabetes. N Engl J Med 373, 2117-2128.

11. Mahaffey, K.W., Neal, B., Perkovic, V., de Zeeuw, D., Fulcher, G., Erondu, N., Shaw, W., Fabbrini, E., Sun, T., Li, Q., et al. (2018). Canagliflozin for Primary and Secondary Prevention of Cardiovascular Events: Results From the CANVAS Program (Canagliflozin Cardiovascular Assessment Study). Circulation 137, 323–334.

12. Wiviott, S.D., Raz, I., Bonaca, M.P., Mosenzon, O., Kato, E.T., Cahn, A., Silverman, M.G., Zelniker, T.A., Kuder, J.F., Murphy, S.A., et al. (2019). Dapagliflozin and Cardiovascular Outcomes in Type 2 Diabetes. N Engl J Med 380, 347–357.

13. Joseph, J., Liu, C., Hui, Q., Aragam, K., Wang, Z., Charest, B., Huffman, J.E., Keaton, J.M., Edwards, T.L., Demissie, S., et al. (2022). Genetic architecture of heart failure with preserved versus reduced ejection fraction. Nat Commun 13, 7753.

14. Levin, M.G., Tsao, N.L., Singhal, P., Liu, C., Vy, H.M.T., Paranjpe, I., Backman, J.D., Bellomo, T.R., Bone, W.P., Biddinger, K.J., et al. (2022). Genome-wide association and multi-trait analyses characterize the common genetic architecture of heart failure. Nat Commun 13, 6914.

15. Shah, S., Henry, A., Roselli, C., Lin, H., Sveinbjornsson, G., Fatemifar, G., Hedman, A.K., Wilk, J.B., Morley, M.P., Chaffin, M.D., et al. (2020). Genome-wide association and Mendelian randomisation analysis provide insights into the pathogenesis of heart failure. Nat Commun 11, 163.

16. Lindgren, M.P., PirouziFard, M., Smith, J.G., Sundquist, J., Sundquist, K., and Zoller, B. (2018). A Swedish Nationwide Adoption Study of the Heritability of Heart Failure. JAMA Cardiol 3, 703–710.

17. Kim, J., Jensen, A., Ko, S., Raghavan, S., Phillips, L.S., Hung, A., Sun, Y., Zhou, H., Reaven, P., and Zhou, J.J. (2022). Systematic Heritability and Heritability Enrichment Analysis for Diabetes Complications in UK Biobank and ACCORD Studies. Diabetes 71, 1137–1148.

18. Zheng, J., Baird, D., Borges, M.C., Bowden, J., Hemani, G., Haycock, P., Evans, D.M., and Smith, G.D. (2017). Recent Developments in Mendelian Randomization Studies. Curr Epidemiol Rep 4, 330–345.

19. Gaziano, J.M., Concato, J., Brophy, M., Fiore, L., Pyarajan, S., Breeling, J., Whitbourne, S., Deen, J., Shannon, C., Humphries, D., et al. (2016). Million Veteran Program: A mega-biobank to study genetic influences on health and disease. J Clin Epidemiol 70, 214–223.

20. Bycroft, C., Freeman, C., Petkova, D., Band, G., Elliott, L.T., Sharp, K., Motyer, A., Vukcevic, D., Delaneau, O., O’Connell, J., et al. (2018). The UK Biobank resource with deep phenotyping and genomic data. Nature 562, 203–209.

21. Kurgansky, K.E., Schubert, P., Parker, R., Djousse, L., Riebman, J.B., Gagnon, D.R., and Joseph, J. (2020). Association of pulse rate with outcomes in heart failure with reduced ejection fraction: a retrospective cohort study. BMC Cardiovasc Disord 20, 92.

22. Patel, Y.R., Robbins, J.M., Kurgansky, K.E., Imran, T., Orkaby, A.R., McLean, R.R., Ho, Y.L., Cho, K., Michael Gaziano, J., Djousse, L., et al. (2018). Development and validation of a heart failure with preserved ejection fraction cohort using electronic medical records. BMC Cardiovasc Disord 18, 128.

23. Rhee, M.K., Ho, Y.L., Raghavan, S., Vassy, J.L., Cho, K., Gagnon, D., Staimez, L.R., Ford, C.N., Wilson, P.W.F., and Phillips, L.S. (2019). Random plasma glucose predicts the diagnosis of diabetes. PLoS One 14, e0219964.

24. Aragam, K.G., Chaffin, M., Levinson, R.T., McDermott, G., Choi, S.H., Shoemaker, M.B., Haas, M.E., Weng, L.C., Lindsay, M.E., Smith, J.G., et al. (2018). Phenotypic Refinement of Heart Failure in a National Biobank Facilitates Genetic Discovery. Circulation.

25. Zhong, H., Magee, M.J., Huang, Y., Hui, Q., Gwinn, M., Gandhi, N.R., and Sun, Y.V. (2020). Evaluation of the Host Genetic Effects of Tuberculosis-Associated Variants Among Patients With Type 1 and Type 2 Diabetes Mellitus. Open Forum Infect Dis 7, ofaa106.

26. Hunter-Zinck, H., Shi, Y., Li, M., Gorman, B.R., Ji, S.G., Sun, N., Webster, T., Liem, A., Hsieh, P., Devineni, P., et al. (2020). Genotyping Array Design and Data Quality Control in the Million Veteran Program. Am J Hum Genet 106, 535–548.

27. Manichaikul, A., Mychaleckyj, J.C., Rich, S.S., Daly, K., Sale, M., and Chen, W.M. (2010). Robust relationship inference in genome-wide association studies. Bioinformatics 26, 2867–2873.

28. Loh, P.R., Palamara, P.F., and Price, A.L. (2016). Fast and accurate long-range phasing in a UK Biobank cohort. Nat Genet 48, 811–816.

29. Das, S., Forer, L., Schonherr, S., Sidore, C., Locke, A.E., Kwong, A., Vrieze, S.I., Chew, E.Y., Levy, S., McGue, M., et al. (2016). Next-generation genotype imputation service and methods. Nat Genet 48, 1284–1287.

30. Fang, H., Hui, Q., Lynch, J., Honerlaw, J., Assimes, T.L., Huang, J., Vujkovic, M., Damrauer, S.M., Pyarajan, S., Gaziano, J.M., et al. (2019). Harmonizing Genetic Ancestry and Self-identified Race/Ethnicity in Genome-wide Association Studies. Am J Hum Genet 105, 763–772.

31. Kurki, M.I., Karjalainen, J., Palta, P., Sipila, T.P., Kristiansson, K., Donner, K.M., Reeve, M.P., Laivuori, H., Aavikko, M., Kaunisto, M.A., et al. (2023). FinnGen provides genetic insights from a well-phenotyped isolated population. Nature 613, 508–518.

32. Magi, R., and Morris, A.P. (2010). GWAMA: software for genome-wide association meta-analysis. BMC Bioinformatics 11, 288.

33. Watanabe, K., Taskesen, E., van Bochoven, A., and Posthuma, D. (2017). Functional mapping and annotation of genetic associations with FUMA. Nat Commun 8, 1826.

34. Scott, R.A., Scott, L.J., Magi, R., Marullo, L., Gaulton, K.J., Kaakinen, M., Pervjakova, N., Pers, T.H., Johnson, A.D., Eicher, J.D., et al. (2017). An Expanded Genome-Wide Association Study of Type 2 Diabetes in Europeans. Diabetes 66, 2888–2902.

35. Verbanck, M., Chen, C.Y., Neale, B., and Do, R. (2018). Detection of widespread horizontal pleiotropy in causal relationships inferred from Mendelian randomization between complex traits and diseases. Nat Genet 50, 693–698.

36. Darrous, L., Mounier, N., and Kutalik, Z. (2021). Simultaneous estimation of bi-directional causal effects and heritable confounding from GWAS summary statistics. Nat Commun 12, 7274.

37. Mahmoud, O., Dudbridge, F., Davey Smith, G., Munafo, M., and Tilling, K. (2022). A robust method for collider bias correction in conditional genome-wide association studies. Nat Commun 13, 619.

38. Johansson, I., Edner, M., Dahlstrom, U., Nasman, P., Ryden, L., and Norhammar, A. (2014). Is the prognosis in patients with diabetes and heart failure a matter of unsatisfactory management? An observational study from the Swedish Heart Failure Registry. Eur J Heart Fail 16, 409–418.

39. Heidenreich, P.A., Bozkurt, B., Aguilar, D., Allen, L.A., Byun, J.J., Colvin, M.M., Deswal, A., Drazner, M.H., Dunlay, S.M., Evers, L.R., et al. (2022). 2022 AHA/ACC/HFSA Guideline for the Management of Heart Failure: Executive Summary: A Report of the American College of Cardiology/American Heart Association Joint Committee on Clinical Practice Guidelines. Circulation 145, e876-e894.

40. Ahlqvist, E., Storm, P., Karajamaki, A., Martinell, M., Dorkhan, M., Carlsson, A., Vikman, P., Prasad, R.B., Aly, D.M., Almgren, P., et al. (2018). Novel subgroups of adult-onset diabetes and their association with outcomes: a data-driven cluster analysis of six variables. Lancet Diabetes Endocrinol 6, 361–369.

41. Hall, H., Perelman, D., Breschi, A., Limcaoco, P., Kellogg, R., McLaughlin, T., and Snyder, M. (2018). Glucotypes reveal new patterns of glucose dysregulation. PLoS Biol 16, e2005143.

42. Udler, M.S., Kim, J., von Grotthuss, M., Bonas-Guarch, S., Cole, J.B., Chiou, J., Christopher, D.A.o.b.o.M., the, I., Boehnke, M., Laakso, M., et al. (2018). Type 2 diabetes genetic loci informed by multi-trait associations point to disease mechanisms and subtypes: A soft clustering analysis. PLoS Med 15, e1002654.

43. Shah, S.J., Katz, D.H., Selvaraj, S., Burke, M.A., Yancy, C.W., Gheorghiade, M., Bonow, R.O., Huang, C.C., and Deo, R.C. (2015). Phenomapping for novel classification of heart failure with preserved ejection fraction. Circulation 131, 269–279.

