## Supplemental Materials, Figures, and Tables for "Correction for Collider Bias in the Genome-wide Association Study of Diabetes-Related Heart Failure due to Bidirectional Relationship between Heart Failure and Type 2 Diabetes"

**Corresponding Author:**

Yan V. Sun, PhD

**Supplementary Information**

**Supplementary Figures**

**Supplementary Figure 1.** Regional plots of chromosome 4 (1A, *PITX2*) and chromosome 9 (1B, *CDKN2B−AS1*) regions associated with diabetes-related HF.

1A

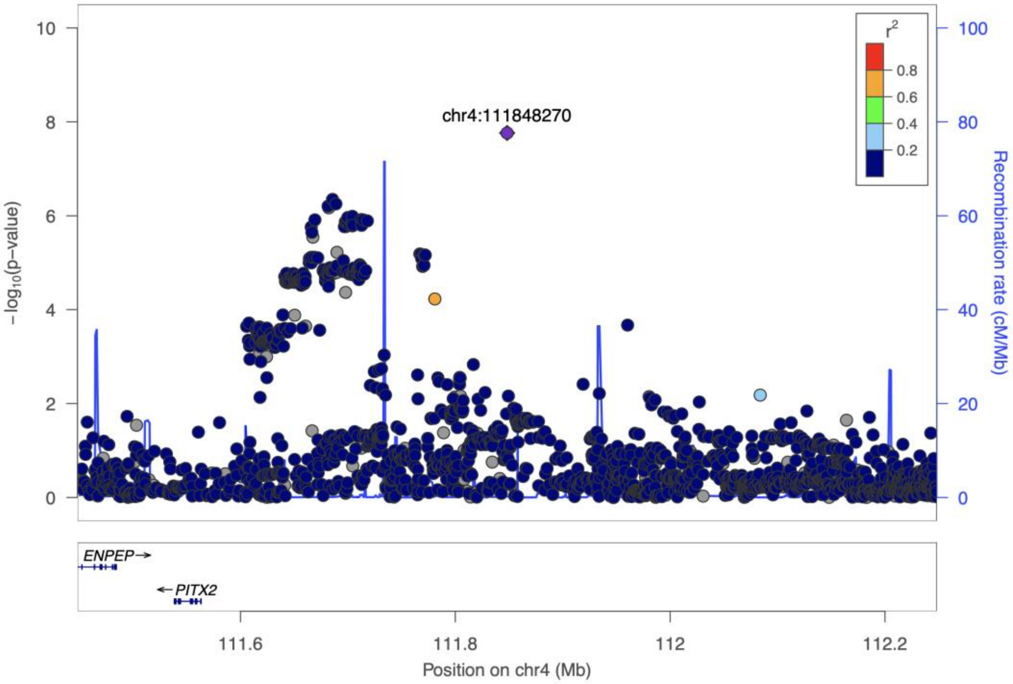


1B

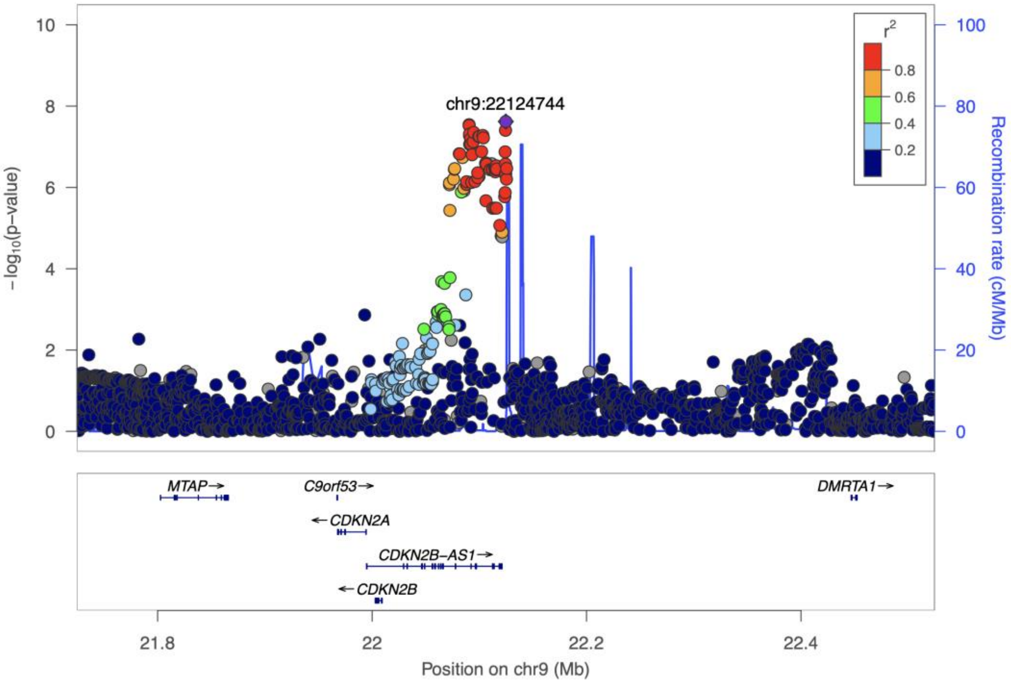

**Supplementary Figure 2.** GWAS of T2D-adjusted all-cause HF (A: Manhattan plot; B: QQ-plot, inflation factor=1.14)

2A

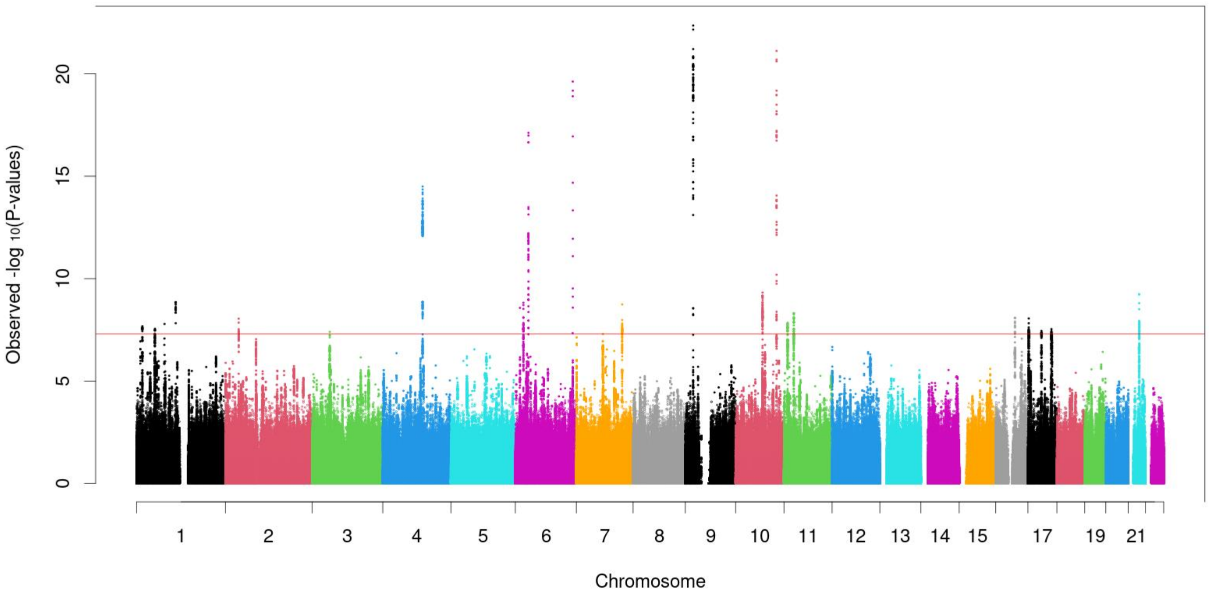


2B

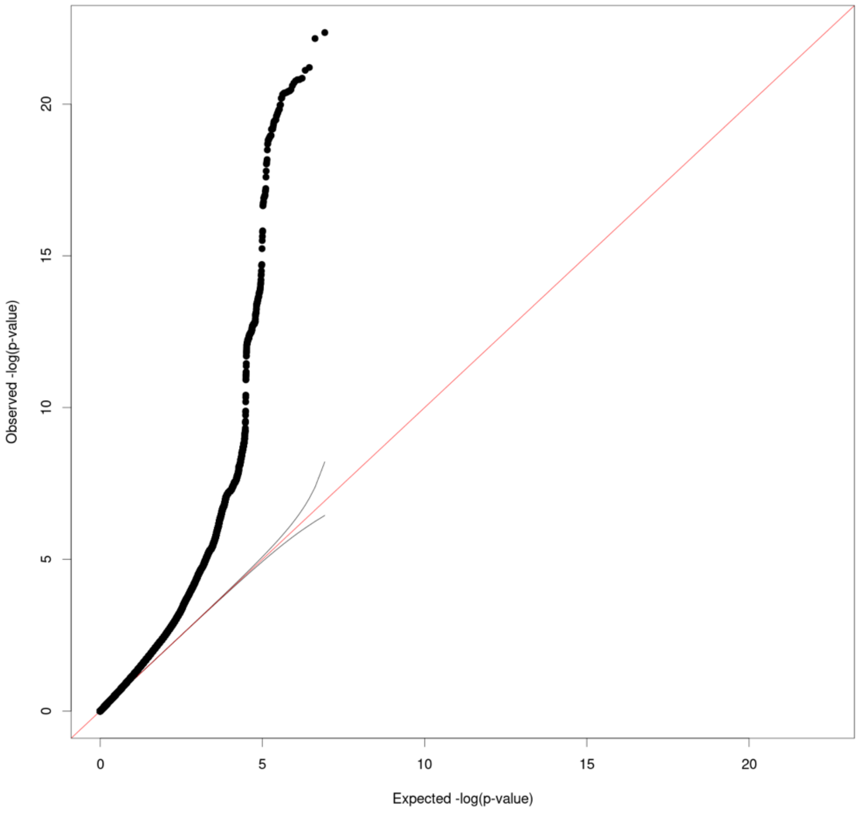

### Supplementary Tables

**Supplementary Table 1**. Characteristics of demographic variables and comorbidities in the UK Biobank cohort.

| | Overall (N=337,423) | | | T2D (N=26,431) | | | Non-T2D (N=310,992) | | |
|---|---|---|---|---|---|---|---|---|---|
| | **HF (N=12191)** | **Non-HF controls (N=325232)** | **P** | **HF (N=3506)** | **Non-HF controls (N=22925)** | **P** | **HF (N=8685)** | **Non-HF controls (N=302307)** | **P** |
| **Age, years (SD)** | 62.11 (6.11) | 56.68 (7.99) | **<0.001** | 62.30 (6.01) | 59.78 (6.97) | **<0.001** | 62.03 (6.15) | 56.45 (8.01) | **<0.001** |
| **Male, n (%)** | 7879 (64.6) | 148341 (45.6) | **<0.001** | 2388 (68.1) | 13578 (59.2) | **<0.001** | 5491 (63.2) | 134763 (44.6) | **<0.001** |
| **BMI, kg/m$^2$ (SD)** | 29.75 (5.70) | 27.31 (4.69) | **<0.001** | 32.68 (6.07) | 31.42 (5.63) | **<0.001** | 28.58 (5.10) | 27.00 (4.46) | **<0.001** |
| **Obesity, (BMI≥30 kg/m$^2$)** | 5048 (41.8) | 76086 (23.5) | **<0.001** | 2199 (63.7) | 12566 (55.2) | **<0.001** | 2849 (33.0) | 63520 (21.1) | **<0.001** |
| **Atrial fibrillation, n (%)** | 1762 (14.5) | 3924 (1.2) | **<0.001** | 563 (16.1) | 606 (2.6) | **<0.001** | 1199 (13.8) | 3318 (1.1) | **<0.001** |
| **Coronary artery disease, n (%)** | 3067 (25.2) | 10559 (3.2) | **<0.001** | 1235 (35.2) | 2504 (10.9) | **<0.001** | 1832 (21.1) | 8055 (2.7) | **<0.001** |
| **Chronic kidney disease, n (%)** | 5161 (44.6) | 93200 (30.1) | **<0.001** | 1565 (47.2) | 7312 (33.6) | **<0.001** | 3596 (43.5) | 85888 (29.8) | **<0.001** |
| **Type 2 Diabetes, n (%)** | 3506 (28.8) | 22925 (7.0) | **<0.001** | - | - | - | - | - | - |
| **Hyperlipidemia, n (%)** | 10114 (85.2) | 249168 (79.7) | **<0.001** | 3050 (88.3) | 19304 (86.2) | **0.001** | 7064 (83.9) | 229864 (79.2) | **<0.001** |
| **Hypertension, n (%)** | 7233 (59.5) | 89938 (27.7) | **<0.001** | 2666 (76.1) | 13915 (60.8) | **<0.001** | 4567 (52.7) | 76023 (25.2) | **<0.001** |

Note: HF: Heart Failure; T2D: Type 2 Diabetes; BMI: Body Mass Index.

**Supplementary Table 2**. The summary statistics of the sentinel SNPs from 61 genome-wide significant loci for all-cause heart failure.

| rsID | Gene | Chr. | Pos. (hg19) | EA | NEA | EAF | OR (95% CI) | P | N |
|---|---|---|---|---|---|---|---|---|---|
| rs1739833 | *C1orf64* | 1 | 16331108 | C | T | 0.670531 | 1.04 (1.03, 1.06) | 8.50E-16 | 1229817 |
| rs28416760 | *INPP5B* | 1 | 38409112 | T | A | 0.733491 | 1.03 (1.02, 1.04) | 1.40E-08 | 1593784 |
| rs78256308 | *DMRTA2* | 1 | 50814474 | G | T | 0.020645 | 1.14 (1.1, 1.18) | 1.60E-14 | 1595050 |
| rs72664353 | *PPAP2B* | 1 | 57001898 | C | T | 0.907617 | 1.05 (1.03, 1.07) | 3.32E-09 | 1591701 |
| rs2474372 | *NFIA* | 1 | 61882251 | A | G | 0.33984 | 1.04 (1.03, 1.05) | 3.23E-12 | 1597477 |
| rs602633 | *PSRC1* | 1 | 109821511 | G | T | 0.784287 | 1.05 (1.04, 1.06) | 6.75E-17 | 1594488 |
| rs11102694 | *BCL2L15* | 1 | 114426001 | A | G | 0.200278 | 1.04 (1.02, 1.05) | 3.06E-09 | 1599523 |
| rs17163313 | *MIA3* | 1 | 222799625 | G | T | 0.712736 | 1.03 (1.02, 1.04) | 3.54E-08 | 1568654 |
| rs12992672 | - | 2 | 632592 | A | G | 0.821402 | 1.04 (1.03, 1.06) | 1.09E-10 | 1595472 |
| rs7595697 | *E2F6* | 2 | 11568158 | T | C | 0.383298 | 1.03 (1.02, 1.04) | 1.94E-09 | 1602778 |
| rs17038861 | *HEATR5B* | 2 | 37233265 | T | G | 0.799884 | 1.04 (1.03, 1.06) | 1.09E-12 | 1604806 |
| rs7564469 | *ZEB2* | 2 | 145258445 | C | T | 0.164794 | 1.04 (1.03, 1.05) | 2.57E-09 | 1602624 |
| rs3820888 | *SPATS2L* | 2 | 201180023 | C | T | 0.403585 | 1.03 (1.02, 1.04) | 1.43E-10 | 1599565 |
| rs6796042 | *FOXP1* | 3 | 71530120 | A | G | 0.621141 | 1.03 (1.02, 1.04) | 7.35E-09 | 1604807 |
| rs10938398 | - | 4 | 45186139 | A | G | 0.435524 | 1.03 (1.02, 1.04) | 1.49E-09 | 1597484 |
| rs17253722 | *SHROOM3* | 4 | 77367287 | G | A | 0.573241 | 1.03 (1.02, 1.04) | 4.57E-08 | 1604785 |
| rs1906618 | *PITX2* | 4 | 111695422 | G | A | 0.124757 | 1.10 (1.09, 1.12) | 2.51E-40 | 1602799 |
| rs17620390 | *CAMK2D* | 4 | 114384328 | C | A | 0.265095 | 1.04 (1.03, 1.05) | 4.08E-11 | 1598150 |
| rs6842241 | *EDNRA* | 4 | 148400819 | A | C | 0.138262 | 1.04 (1.02, 1.05) | 4.67E-08 | 1596428 |
| rs11746435 | *KLHL3* | 5 | 137006762 | A | T | 0.769082 | 1.04 (1.03, 1.05) | 1.08E-10 | 1597234 |
| rs72810976 | *CPEB4* | 5 | 173309057 | G | A | 0.680527 | 1.03 (1.02, 1.04) | 1.05E-08 | 1600502 |
| rs6909574 | *HDGFL1* | 6 | 22606773 | G | A | 0.331273 | 1.04 (1.03, 1.05) | 5.99E-14 | 1596469 |
| rs3176326 | *CDKN1A* | 6 | 36647289 | G | A | 0.809434 | 1.08 (1.06, 1.09) | 1.71E-31 | 1597228 |
| rs9443648 | *PHIP* | 6 | 79853605 | A | G | 0.464011 | 1.03 (1.02, 1.04) | 2.79E-09 | 1604777 |

| | | | | | | | | | |
|---|---|---|---|---|---|---|---|---|---|
| rs117321970 | *FHL5* | 6 | 97071980 | T | C | 0.045267 | 1.07 (1.05, 1.10) | 2.81E-08 | 1577505 |
| rs10455872 | *LPA* | 6 | 161010118 | G | A | 0.063917 | 1.1 (1.08, 1.12) | 7.11E-24 | 1602834 |
| rs35005436 | *GTF2I* | 7 | 74134911 | C | T | 0.148818 | 1.05 (1.03, 1.06) | 1.45E-11 | 1575530 |
| rs6945340 | *POM121C* | 7 | 75100124 | C | T | 0.215821 | 1.04 (1.03, 1.05) | 6.74E-11 | 1550057 |
| rs11773884 | *CDK6* | 7 | 92285123 | A | G | 0.693838 | 1.03 (1.02, 1.04) | 9.80E-10 | 1607789 |
| rs3918226 | *NOS3* | 7 | 150690176 | T | C | 0.079657 | 1.05 (1.03, 1.07) | 3.54E-08 | 1594659 |
| rs4733328 | *NRG1* | 8 | 32259246 | G | A | 0.142002 | 1.04 (1.03, 1.05) | 3.44E-08 | 1602596 |
| rs11774829 | *RP11-127H5.1* | 8 | 105978368 | T | A | 0.880685 | 1.05 (1.03, 1.07) | 3.76E-10 | 1605769 |
| rs1537371 | *RP11-145E5.5* | 9 | 22099568 | A | C | 0.474101 | 1.06 (1.05, 1.07) | 4.48E-32 | 1607858 |
| rs7873569 | *TMEM245* | 9 | 111796753 | A | T | 0.569484 | 1.03 (1.02, 1.04) | 1.61E-08 | 1607875 |
| rs600038 | - | 9 | 136151806 | C | T | 0.21379 | 1.05 (1.04, 1.06) | 1.26E-17 | 1602628 |
| rs60212594 | *SYNPO2L* | 10 | 75414344 | G | C | 0.85448 | 1.06 (1.04, 1.07) | 4.31E-16 | 1602630 |
| rs17617337 | *BAG3* | 10 | 121426884 | C | T | 0.781435 | 1.05 (1.03, 1.06) | 8.89E-15 | 1607844 |
| rs71311904 | *BDNF* | 11 | 27742447 | C | CCATTT | 0.815465 | 1.05 (1.03, 1.06) | 3.32E-09 | 643819 |
| rs4755720 | *HSD17B12* | 11 | 43628749 | C | T | 0.38746 | 1.04 (1.02, 1.05) | 5.21E-12 | 1597510 |
| rs113104597 | *CHD4* | 12 | 6703172 | C | T | 0.162382 | 1.04 (1.03, 1.05) | 1.21E-08 | 1232913 |
| rs34682944 | *DIP2B* | 12 | 50982864 | A | G | 0.311432 | 1.03 (1.02, 1.05) | 1.07E-08 | 818352 |
| rs2013002 | *RP11-162P23.2* | 12 | 112200150 | T | C | 0.406842 | 1.03 (1.02, 1.04) | 3.14E-11 | 1595137 |
| rs112403212 | *SCARB1* | 12 | 125303254 | T | C | 0.140563 | 1.05 (1.03, 1.06) | 7.99E-09 | 1259036 |
| rs10161594 | *ATP4B* | 13 | 114306243 | G | C | 0.139972 | 1.04 (1.03, 1.06) | 1.09E-08 | 1597800 |
| rs10149845 | *PRKD1* | 14 | 30177079 | T | C | 0.415774 | 1.03 (1.02, 1.04) | 2.46E-09 | 1602586 |
| rs58472533 | *AMN* | 14 | 103385634 | G | A | 0.203113 | 1.04 (1.03, 1.05) | 5.49E-10 | 1602421 |
| rs17483686 | *IREB2* | 15 | 78733390 | T | A | 0.329609 | 1.03 (1.02, 1.04) | 1.55E-09 | 1234895 |
| rs11634851 | *ABHD17C* | 15 | 81028965 | G | C | 0.454433 | 1.03 (1.02, 1.04) | 4.87E-08 | 1602537 |
| rs11642015 | *FTO* | 16 | 53802494 | T | C | 0.413919 | 1.05 (1.04, 1.06) | 1.78E-26 | 1600562 |
| rs12929503 | *NFAT5* | 16 | 69565461 | T | C | 0.577005 | 1.03 (1.02, 1.05) | 1.10E-09 | 1259029 |
| rs2106261 | *ZFHX3* | 16 | 73051620 | T | C | 0.185836 | 1.03 (1.02, 1.05) | 2.96E-08 | 1597551 |
| rs8046000 | *CFDP1* | 16 | 75433883 | G | C | 0.583977 | 1.03 (1.02, 1.04) | 2.35E-10 | 1602603 |

| | | | | | | | | | |
|---|---|---|---|---|---|---|---|---|---|
| rs11861290 | *CMIP* | 16 | 81548522 | A | G | 0.76268 | 1.04 (1.03, 1.05) | 2.27E-10 | 1605856 |
| rs12950555 | *SMG6* | 17 | 2156910 | C | G | 0.351885 | 1.04 (1.03, 1.05) | 5.87E-15 | 1234894 |
| rs11656489 | *ADORA2B* | 17 | 15837141 | G | C | 0.193086 | 1.04 (1.02, 1.05) | 6.15E-09 | 1605860 |
| rs11658278 | *ZPBP2* | 17 | 38031164 | T | C | 0.479866 | 1.03 (1.02, 1.04) | 1.73E-08 | 1602622 |
| rs17608766 | *GOSR2; RP11-156P1.2* | 17 | 45013271 | C | T | 0.146188 | 1.04 (1.03, 1.06) | 6.24E-10 | 1599531 |
| rs113437066 | *BPTF* | 17 | 65836220 | A | ATTT | 0.197489 | 1.06 (1.04, 1.08) | 1.87E-10 | 302258 |
| rs1788826 | *NPC1* | 18 | 21154024 | G | A | 0.358788 | 1.03 (1.02, 1.04) | 8.22E-10 | 1605877 |
| rs4327120 | - | 18 | 36532976 | T | C | 0.897987 | 1.05 (1.03, 1.07) | 7.00E-10 | 1599531 |
| rs2832275 | *BACH1* | 21 | 30602994 | T | A | 0.143356 | 1.05 (1.03, 1.06) | 5.61E-11 | 1605855 |

Gene: gene abbreviation of the gene closest to the sentinel SNP; Chr.: Chromosome; Pos.: Position; EA: Effect Allele (i.e., risk allele for heart failure); NEA: Non-effect Allele; EAF: Effect Allele Frequency; OR: Odds Ratio; P: P value; N: sample size.

**Supplementary Table 3**. Summary statistics of 59 independent genetic instruments (SNPs) strongly associated with all-cause heart failure (meta-analysis p-value<$5\times10^{-8}$).

| rsID | Gene | Chr. | Pos. (hg19) | EA | NEA | Associations with HF | | | | | Associations with T2D | | | |
|---|---|---|---|---|---|---|---|---|---|---|---|---|---|---|
| | | | | | | **EAF** | **BETA** | **SE** | **P** | **N** | **BETA** | **SE** | **P** | **N** |
| rs1739833 | *C1orf64* | 1 | 16331108 | C | T | 0.671 | 0.044 | 0.005 | 8.50E-16 | 1229817 | 0.017 | 0.013 | 0.17 | 158184 |
| rs28416760 | *INPP5B* | 1 | 38409112 | T | A | 0.733 | 0.032 | 0.006 | 1.40E-08 | 1593784 | 0.008 | 0.014 | 0.58 | 158184 |
| rs78256308 | *DMRTA2* | 1 | 50814474 | G | T | 0.021 | 0.133 | 0.017 | 1.60E-14 | 1595050 | 0.043 | 0.043 | 0.31 | 158184 |
| rs72664353 | *PPAP2B* | 1 | 57001898 | C | T | 0.908 | 0.051 | 0.009 | 3.32E-09 | 1591701 | 0.03 | 0.021 | 0.15 | 158184 |
| rs2474372 | *NFIA* | 1 | 61882251 | A | G | 0.34 | 0.036 | 0.005 | 3.23E-12 | 1597477 | 0.015 | 0.013 | 0.24 | 158184 |
| rs602633 | *PSRC1* | 1 | 109821511 | G | T | 0.784 | 0.049 | 0.006 | 6.75E-17 | 1594488 | -0.026 | 0.015 | 0.081 | 158184 |
| rs11102694 | *BCL2L15* | 1 | 114426001 | A | G | 0.2 | 0.036 | 0.006 | 3.06E-09 | 1599523 | -0.005 | 0.016 | 0.77 | 158184 |
| rs17163313 | *MIA3* | 1 | 222799625 | G | T | 0.713 | 0.03 | 0.005 | 3.54E-08 | 1568654 | 0.013 | 0.014 | 0.35 | 150161 |
| rs12992672 | - | 2 | 632592 | A | G | 0.821 | 0.042 | 0.007 | 1.09E-10 | 1595472 | 0.044 | 0.015 | 0.0043 | 158186 |
| rs7595697 | *E2F6* | 2 | 11568158 | T | C | 0.383 | 0.029 | 0.005 | 1.94E-09 | 1602778 | 0 | 0.012 | 0.99 | 158186 |
| rs17038861 | *HEATR5B* | 2 | 37233265 | T | G | 0.8 | 0.043 | 0.006 | 1.09E-12 | 1604806 | 0.016 | 0.016 | 0.32 | 152599 |
| rs7564469 | *ZEB2* | 2 | 145258445 | C | T | 0.165 | 0.038 | 0.006 | 2.57E-09 | 1602624 | 0.001 | 0.017 | 0.96 | 152599 |
| rs3820888 | *SPATS2L* | 2 | 201180023 | C | T | 0.404 | 0.032 | 0.005 | 1.43E-10 | 1599565 | 0.017 | 0.012 | 0.16 | 158186 |
| rs6796042 | *FOXP1* | 3 | 71530120 | A | G | 0.621 | 0.029 | 0.005 | 7.35E-09 | 1604807 | 0.012 | 0.012 | 0.33 | 158185 |
| rs10938398 | - | 4 | 45186139 | A | G | 0.436 | 0.03 | 0.005 | 1.49E-09 | 1597484 | 0.04 | 0.012 | 0.0012 | 158185 |
| rs17253722 | *SHROOM3* | 4 | 77367287 | G | A | 0.573 | 0.027 | 0.005 | 4.57E-08 | 1604785 | -0.001 | 0.013 | 0.97 | 152598 |
| rs1906618 | *PITX2* | 4 | 111695422 | G | A | 0.125 | 0.096 | 0.007 | 2.51E-40 | 1602799 | 0.041 | 0.019 | 0.028 | 158185 |
| rs17620390 | *CAMK2D* | 4 | 114384328 | C | A | 0.265 | 0.036 | 0.005 | 4.08E-11 | 1598150 | 0.023 | 0.014 | 0.093 | 152598 |
| rs6842241 | *EDNRA* | 4 | 148400819 | A | C | 0.138 | 0.038 | 0.007 | 4.67E-08 | 1596428 | 0.006 | 0.017 | 0.73 | 158185 |
| rs11746435 | *KLHL3* | 5 | 137006762 | A | T | 0.769 | 0.037 | 0.006 | 1.08E-10 | 1597234 | 0.009 | 0.014 | 0.51 | 158186 |
| rs72810976 | *CPEB4* | 5 | 173309057 | G | A | 0.681 | 0.03 | 0.005 | 1.05E-08 | 1600502 | -0.007 | 0.013 | 0.59 | 158186 |
| rs6909574 | *HDGFL1* | 6 | 22606773 | G | A | 0.331 | 0.038 | 0.005 | 5.99E-14 | 1596469 | -0.001 | 0.013 | 0.95 | 158185 |

| | | | | | | | | | | | | | | |
|---|---|---|---|---|---|---|---|---|---|---|---|---|---|---|
| rs3176326 | *CDKN1A* | 6 | 36647289 | G | A | 0.809 | 0.073 | 0.006 | 1.71E-31 | 1597228 | 0.031 | 0.016 | 0.045 | 158185 |
| rs9443648 | *PHIP* | 6 | 79853605 | A | G | 0.464 | 0.029 | 0.005 | 2.79E-09 | 1604777 | 0.017 | 0.013 | 0.17 | 152598 |
| rs117321970 | *FHL5* | 6 | 97071980 | T | C | 0.045 | 0.07 | 0.013 | 2.81E-08 | 1577505 | 0.005 | 0.035 | 0.89 | 158185 |
| rs10455872 | *LPA* | 6 | 161010118 | G | A | 0.064 | 0.098 | 0.01 | 7.11E-24 | 1602834 | 0.013 | 0.028 | 0.63 | 158185 |
| rs35005436 | *GTF2I* | 7 | 74134911 | C | T | 0.149 | 0.048 | 0.007 | 1.45E-11 | 1575530 | 0.029 | 0.019 | 0.13 | 142913 |
| rs6945340 | *POM121C* | 7 | 75100124 | C | T | 0.216 | 0.04 | 0.006 | 6.74E-11 | 1550057 | 0.004 | 0.018 | 0.84 | 138337 |
| rs11773884 | *CDK6* | 7 | 92285123 | A | G | 0.694 | 0.032 | 0.005 | 9.80E-10 | 1607789 | 0.019 | 0.013 | 0.15 | 158184 |
| rs3918226 | *NOS3* | 7 | 150690176 | T | C | 0.08 | 0.051 | 0.009 | 3.54E-08 | 1594659 | -0.018 | 0.025 | 0.46 | 158184 |
| rs4733328 | *NRG1* | 8 | 32259246 | G | A | 0.142 | 0.039 | 0.007 | 3.44E-08 | 1602596 | 0.002 | 0.018 | 0.93 | 158185 |
| rs11774829 | *RP11-127H5.1* | 8 | 105978368 | T | A | 0.881 | 0.049 | 0.008 | 3.76E-10 | 1605769 | 0.059 | 0.02 | 0.0031 | 152598 |
| rs1537371 | *RP11-145E5.5* | 9 | 22099568 | A | C | 0.474 | 0.057 | 0.005 | 4.48E-32 | 1607858 | 0.037 | 0.012 | 0.0024 | 158183 |
| rs7873569 | *TMEM245* | 9 | 111796753 | A | T | 0.569 | 0.028 | 0.005 | 1.61E-08 | 1607875 | -0.001 | 0.012 | 0.94 | 158183 |
| rs600038* | - | 9 | 136151806 | C | T | 0.214 | 0.05 | 0.006 | 1.26E-17 | 1602628 | 0.074 | 0.015 | 1.00E-06 | 158183 |
| rs60212594 | *SYNPO2L* | 10 | 75414344 | G | C | 0.854 | 0.056 | 0.007 | 4.31E-16 | 1602630 | 0.004 | 0.017 | 0.83 | 158186 |
| rs17617337 | *BAG3* | 10 | 121426884 | C | T | 0.781 | 0.046 | 0.006 | 8.89E-15 | 1607844 | 0.002 | 0.015 | 0.88 | 158186 |
| rs4755720 | *HSD17B12* | 11 | 43628749 | C | T | 0.387 | 0.034 | 0.005 | 5.21E-12 | 1597510 | 0.029 | 0.012 | 0.022 | 158185 |
| rs113104597 | *CHD4* | 12 | 6703172 | C | T | 0.162 | 0.039 | 0.007 | 1.21E-08 | 1232913 | 0.055 | 0.017 | 0.0013 | 158183 |
| rs34682944 | *DIP2B* | 12 | 50982864 | A | G | 0.311 | 0.033 | 0.006 | 1.07E-08 | 818352 | 0.001 | 0.013 | 0.95 | 158183 |
| rs2013002 | *RP11-162P23.2* | 12 | 112200150 | T | C | 0.407 | 0.033 | 0.005 | 3.14E-11 | 1595137 | 0.017 | 0.013 | 0.18 | 158183 |
| rs112403212 | *SCARB1* | 12 | 125303254 | T | C | 0.141 | 0.046 | 0.008 | 7.99E-09 | 1259036 | -0.005 | 0.018 | 0.8 | 158183 |
| rs10161594 | *ATP4B* | 13 | 114306243 | G | C | 0.14 | 0.04 | 0.007 | 1.09E-08 | 1597800 | 0.013 | 0.018 | 0.49 | 158184 |
| rs10149845 | *PRKD1* | 14 | 30177079 | T | C | 0.416 | 0.029 | 0.005 | 2.46E-09 | 1602586 | 0.016 | 0.012 | 0.2 | 158185 |

| | | | | | | | | | | | | | | |
|---|---|---|---|---|---|---|---|---|---|---|---|---|---|---|
| rs58472533 | *AMN* | 14 | 103385634 | G | A | 0.203 | 0.038 | 0.006 | 5.49E-10 | 1602421 | 0.044 | 0.016 | 0.005 | 158185 |
| rs17483686 | *IREB2* | 15 | 78733390 | T | A | 0.33 | 0.032 | 0.005 | 1.55E-09 | 1234895 | -0.011 | 0.013 | 0.37 | 158185 |
| rs11634851 | *ABHD17C* | 15 | 81028965 | G | C | 0.454 | 0.026 | 0.005 | 4.87E-08 | 1602537 | -0.019 | 0.012 | 0.12 | 158185 |
| rs11642015* | *FTO* | 16 | 53802494 | T | C | 0.414 | 0.052 | 0.005 | 1.78E-26 | 1600562 | 0.13 | 0.012 | 5.00E-25 | 158182 |
| rs12929503 | *NFAT5* | 16 | 69565461 | T | C | 0.577 | 0.034 | 0.006 | 1.10E-09 | 1259029 | 0.028 | 0.012 | 0.023 | 158182 |
| rs2106261 | *ZFHX3* | 16 | 73051620 | T | C | 0.186 | 0.034 | 0.006 | 2.96E-08 | 1597551 | -0.002 | 0.016 | 0.9 | 152595 |
| rs8046000 | *CFDP1* | 16 | 75433883 | G | C | 0.584 | 0.031 | 0.005 | 2.35E-10 | 1602603 | 0.002 | 0.012 | 0.86 | 158182 |
| rs11861290 | *CMIP* | 16 | 81548522 | A | G | 0.763 | 0.036 | 0.006 | 2.27E-10 | 1605856 | 0.016 | 0.014 | 0.26 | 158182 |
| rs12950555 | *SMG6* | 17 | 2156910 | C | G | 0.352 | 0.041 | 0.005 | 5.87E-15 | 1234894 | 0 | 0.012 | 0.98 | 158182 |
| rs11656489 | *ADORA2B* | 17 | 15837141 | G | C | 0.193 | 0.035 | 0.006 | 6.15E-09 | 1605860 | -0.013 | 0.016 | 0.41 | 158182 |
| rs11658278 | *ZPBP2* | 17 | 38031164 | T | C | 0.48 | 0.027 | 0.005 | 1.73E-08 | 1602622 | 0.013 | 0.012 | 0.27 | 158182 |
| rs17608766 | *GOSR2; RP11-156P1.2* | 17 | 45013271 | C | T | 0.146 | 0.042 | 0.007 | 6.24E-10 | 1599531 | 0.036 | 0.018 | 0.044 | 158182 |
| rs1788826 | *NPC1* | 18 | 21154024 | G | A | 0.359 | 0.031 | 0.005 | 8.22E-10 | 1605877 | 0.035 | 0.012 | 0.0044 | 158184 |
| rs4327120 | - | 18 | 36532976 | T | C | 0.898 | 0.05 | 0.008 | 7.00E-10 | 1599531 | 0.073 | 0.019 | 0.00016 | 158184 |
| rs2832275 | *BACH1* | 21 | 30602994 | T | A | 0.143 | 0.045 | 0.007 | 5.61E-11 | 1605855 | -0.013 | 0.017 | 0.45 | 158185 |

Gene: gene abbreviation of the gene closest to the sentinel SNP; Chr.: Chromosome; Pos.: Position; HF: Heart Failure; T2D: Type 2 Diabetes; EA: Effect Allele; NEA: Non-effect Allele; EAF: Effect Allele Frequency; BETA: Beta coefficient; SE: Standard Error; P: P value; N: sample size.

*Genetic instruments removed in MR-PRESSO.

**Supplementary Table 4**. Summary statistics of 82 independent genetic instruments (SNPs) strongly associated with T2D from published GWAS.

| rsID | Gene | Chr. | Pos. (hg19) | EA | NEA | Associations with T2D | | | | | Associations with HF | | | | |
|---|---|---|---|---|---|---|---|---|---|---|---|---|---|---|---|
| | | | | | | EAF | BETA | SE | P | N | EAF | BETA | SE | P | N |
| rs3768321 | *MACF1* | 1 | 40035928 | T | G | 0.193 | 0.076 | 0.015 | 8.10E-07 | 158184 | 0.191 | 0.005 | 0.008 | 0.5569 | 1297238 |
| rs12031920 | *FAF1* | 1 | 51109269 | T | A | 0.556 | 0.051 | 0.012 | 3.80E-05 | 158184 | 0.538 | 0.002 | 0.006 | 0.756 | 1291925 |
| rs406767 | *NOTCH2* | 1 | 120554048 | C | T | 0.09 | 0.13 | 0.027 | 7.50E-07 | 136187 | 0.109 | 0.003 | 0.008 | 0.6797 | 1533749 |
| rs340874 | *PROX1* | 1 | 214159256 | C | T | 0.55 | 0.068 | 0.012 | 3.40E-08 | 158184 | 0.527 | 0.002 | 0.005 | 0.6534 | 1594489 |
| rs6757251 | *THADA* | 2 | 43734847 | C | T | 0.901 | 0.13 | 0.021 | 1.90E-10 | 158186 | 0.909 | 0.002 | 0.008 | 0.8337 | 1602833 |
| rs10193447 | *BCL11A* | 2 | 60552476 | T | C | 0.598 | 0.071 | 0.012 | 1.30E-08 | 158186 | 0.599 | 0.009 | 0.005 | 0.0822 | 1599496 |
| rs2972156 | *IRS1* | 2 | 227117778 | G | C | 0.614 | 0.076 | 0.013 | 1.20E-09 | 158186 | 0.635 | 0.008 | 0.006 | 0.1612 | 815252 |
| rs11712037 | *PPARG* | 3 | 12344730 | C | G | 0.875 | 0.13 | 0.019 | 8.60E-13 | 158185 | 0.864 | 0.006 | 0.007 | 0.3655 | 1597573 |
| rs35352848 | *UBE2E2* | 3 | 23455582 | T | C | 0.777 | 0.083 | 0.015 | 1.50E-08 | 158185 | 0.766 | -0.004 | 0.006 | 0.4639 | 1599568 |
| rs7428936 | *ADAMTS9* | 3 | 64710850 | T | C | 0.591 | 0.07 | 0.012 | 1.00E-08 | 158185 | 0.6 | 0.001 | 0.005 | 0.8596 | 1602739 |
| rs11708067 | *ADCY5* | 3 | 123065778 | A | G | 0.787 | 0.11 | 0.015 | 8.80E-13 | 152598 | 0.788 | -0.001 | 0.006 | 0.8873 | 1594489 |
| rs4402960 | *IGF2BP2* | 3 | 185511687 | T | G | 0.306 | 0.14 | 0.013 | 2.70E-25 | 152598 | 0.312 | 0.014 | 0.005 | 0.0068 | 1596469 |
| rs6777684 | *LPP* | 3 | 187741842 | G | A | 0.606 | 0.05 | 0.012 | 5.90E-05 | 158185 | 0.609 | 0.015 | 0.005 | 0.0035 | 1599476 |
| rs3821943 | *WFS1* | 4 | 6299940 | T | C | 0.535 | 0.1 | 0.012 | 4.20E-16 | 158185 | 0.529 | 0.019 | 0.005 | 0.0001 | 1594489 |
| rs7660590 | *TMEM154* | 4 | 153397823 | C | T | 0.716 | 0.054 | 0.014 | 6.80E-05 | 158185 | 0.707 | 0.01 | 0.007 | 0.1484 | 1300518 |
| rs60780116 | *ACSL1* | 4 | 185708807 | T | C | 0.835 | 0.09 | 0.017 | 7.40E-08 | 158185 | 0.827 | -0.002 | 0.006 | 0.7525 | 1597444 |
| rs11747901 | *ARL15* | 5 | 53301561 | G | C | 0.188 | 0.072 | 0.016 | 1.20E-05 | 158186 | 0.243 | -0.008 | 0.006 | 0.2144 | 818352 |
| rs173964 | *ANKRD55* | 5 | 55809465 | G | A | 0.744 | 0.061 | 0.014 | 1.40E-05 | 158186 | 0.724 | 0.003 | 0.007 | 0.6687 | 1298380 |

| | | | | | | | | | | | | | | | |
|---|---|---|---|---|---|---|---|---|---|---|---|---|---|---|---|
| rs9687833 | *ANKRD55* | 5 | 55861601 | A | G | 0.187 | 0.095 | 0.016 | 1.60E-09 | 158186 | 0.185 | 0.013 | 0.006 | 0.031 | 1600557 |
| rs6453287 | *ZBED3* | 5 | 76453765 | C | A | 0.304 | 0.063 | 0.014 | 4.50E-06 | 158186 | 0.274 | 0.001 | 0.005 | 0.8846 | 1602317 |
| rs74944275 | *PAM* | 5 | 102726073 | T | C | 0.041 | 0.15 | 0.033 | 4.40E-06 | 158186 | 0.043 | 0.007 | 0.012 | 0.5648 | 1598068 |
| rs6923241 | *SSR1/RREB1* | 6 | 7258847 | C | T | 0.712 | 0.066 | 0.014 | 1.60E-06 | 158185 | 0.707 | 0.004 | 0.005 | 0.4845 | 1597296 |
| rs7451008 | *CDKAL1* | 6 | 20673880 | C | T | 0.261 | 0.17 | 0.013 | 3.80E-37 | 158185 | 0.279 | 0.011 | 0.005 | 0.0453 | 1599540 |
| rs115321690 | *POU5F1/TCF19* | 6 | 31116526 | G | A | 0.678 | 0.068 | 0.013 | 5.10E-07 | 152598 | 0.694 | 0.012 | 0.005 | 0.0266 | 1594222 |
| rs9271774 | *HLA-DQA1* | 6 | 32594309 | C | A | 0.742 | 0.092 | 0.018 | 3.30E-07 | 135283 | 0.759 | 0.017 | 0.01 | 0.0903 | 474229 |
| rs11759026 | *CENPW* | 6 | 126792095 | G | A | 0.237 | 0.091 | 0.015 | 5.80E-10 | 158185 | 0.236 | 0.001 | 0.007 | 0.8808 | 1295305 |
| rs6918311 | *SLC35D3* | 6 | 137287702 | A | G | 0.528 | 0.069 | 0.014 | 6.70E-07 | 143306 | 0.526 | 0.011 | 0.005 | 0.0223 | 1599538 |
| rs10276674 | *DGKB* | 7 | 14922007 | C | T | 0.198 | 0.085 | 0.016 | 5.10E-08 | 158184 | 0.197 | -0.002 | 0.006 | 0.69 | 1597551 |
| rs10238625 | *DGKB* | 7 | 15054232 | A | G | 0.54 | 0.067 | 0.012 | 3.20E-08 | 158184 | 0.536 | 0.006 | 0.005 | 0.2067 | 1598579 |
| rs1635852 | *JAZF1* | 7 | 28189411 | T | C | 0.502 | 0.092 | 0.012 | 3.00E-14 | 158184 | 0.501 | 0.008 | 0.005 | 0.118 | 1599531 |
| rs10954284 | *KLF14* | 7 | 130463758 | T | A | 0.502 | 0.054 | 0.013 | 1.80E-05 | 152597 | 0.514 | 0.018 | 0.005 | 0.0002 | 1600974 |
| rs1182436 | *MNX1* | 7 | 157027753 | C | T | 0.798 | 0.079 | 0.016 | 8.30E-07 | 156108 | 0.826 | 0.01 | 0.007 | 0.1467 | 1234900 |
| rs516946 | *ANK1* | 8 | 41519248 | C | T | 0.775 | 0.073 | 0.015 | 8.60E-07 | 158185 | 0.775 | 0.006 | 0.006 | 0.3242 | 1597551 |
| rs4734285 | *TP53INP1* | 8 | 95882365 | T | C | 0.62 | 0.055 | 0.013 | 1.50E-05 | 158185 | 0.625 | 0.003 | 0.006 | 0.6441 | 1300178 |
| rs11786613 | *TP53INP1* | 8 | 95957984 | C | A | 0.032 | 0.19 | 0.039 | 1.60E-06 | 155750 | 0.026 | 0.024 | 0.017 | 0.1617 | 1600080 |
| rs3802177 | *SLC30A8* | 8 | 118185025 | G | A | 0.677 | 0.11 | 0.013 | 1.70E-17 | 158185 | 0.67 | -0.001 | 0.005 | 0.7917 | 1593785 |
| rs10965223* | *CDKN2A/B* | 9 | 22067004 | A | G | 0.592 | 0.077 | 0.012 | 4.00E-10 | 158183 | 0.598 | 0.03 | 0.005 | 1.03E-09 | 1602629 |
| rs10965248 | *CDKN2A/B* | 9 | 22132878 | T | C | 0.819 | 0.14 | 0.016 | 6.40E-17 | 158183 | 0.828 | -0.006 | 0.006 | 1.03 | 1598116 |
| rs13301067 | *TLE4* | 9 | 81900744 | G | A | 0.924 | 0.1 | 0.024 | 1.50E-05 | 158183 | 0.93 | -0.001 | 0.011 | 0.9405 | 1261007 |
| rs9410573 | *TLE1* | 9 | 84311800 | T | C | 0.599 | 0.073 | 0.013 | 2.00E-08 | 158183 | 0.594 | 0.001 | 0.005 | 0.8653 | 1602530 |

| | | | | | | | | | | | | | | | |
|---|---|---|---|---|---|---|---|---|---|---|---|---|---|---|---|
| rs635634* | *ABO* | 9 | 136155000 | T | C | 0.18 | 0.081 | 0.016 | 3.60E-07 | 158183 | 0.192 | 0.049 | 0.006 | 1.18E-15 | 1585859 |
| rs11257659 | *CDC123/CAMK1D* | 10 | 12309269 | T | C | 0.229 | 0.081 | 0.015 | 2.70E-08 | 158186 | 0.248 | 0.004 | 0.009 | 0.6537 | 516060 |
| rs810517 | *ZMIZ1* | 10 | 80942620 | C | T | 0.515 | 0.089 | 0.013 | 1.30E-12 | 152599 | 0.525 | 0.005 | 0.005 | 0.2898 | 1605895 |
| rs11187140 | *HHEX/IDE* | 10 | 94466910 | G | A | 0.622 | 0.13 | 0.013 | 4.20E-26 | 158186 | 0.628 | 0 | 0.005 | 0.956 | 1600594 |
| rs7903146* | *TCF7L2* | 10 | 114758349 | T | C | 0.289 | 0.29 | 0.013 | 9.30E-108 | 158186 | 0.271 | -0.002 | 0.005 | 0.6954 | 1597551 |
| rs2292626 | *PLEKHA1* | 10 | 124186714 | C | T | 0.504 | 0.085 | 0.012 | 1.80E-12 | 158186 | 0.471 | -0.006 | 0.006 | 0.3044 | 1305612 |
| rs756852 | *KCNQ1* | 11 | 2663891 | G | A | 0.598 | 0.09 | 0.014 | 3.60E-10 | 158185 | 0.613 | -0.006 | 0.007 | 0.4041 | 1254423 |
| rs231360 | *KCNQ1* | 11 | 2692249 | T | C | 0.407 | 0.079 | 0.013 | 9.50E-10 | 158185 | 0.418 | -0.003 | 0.005 | 0.5961 | 1598812 |
| rs233449 | *KCNQ1* | 11 | 2843803 | G | A | 0.726 | 0.089 | 0.014 | 2.80E-10 | 158185 | 0.738 | 0.008 | 0.005 | 0.1393 | 1596542 |
| rs2237897 | *KCNQ1* | 11 | 2858546 | C | T | 0.946 | 0.22 | 0.031 | 4.90E-13 | 158185 | 0.943 | 0.018 | 0.014 | 0.1782 | 1297357 |
| rs441613 | *KCNQ1* | 11 | 2910191 | C | T | 0.633 | 0.054 | 0.013 | 3.30E-05 | 158185 | 0.632 | -0.006 | 0.005 | 0.2721 | 1602440 |
| rs5219 | *KCNJ11* | 11 | 17409572 | T | C | 0.383 | 0.068 | 0.012 | 4.30E-08 | 158185 | 0.396 | 0.019 | 0.005 | 0.0001 | 1592307 |
| rs1061810* | *HSD17B12* | 11 | 43877934 | A | C | 0.279 | 0.08 | 0.014 | 5.30E-09 | 158185 | 0.289 | 0.031 | 0.005 | 4.58E-09 | 1607805 |
| rs111669836 | *MAP3K11* | 11 | 65364385 | A | T | 0.249 | 0.07 | 0.014 | 7.40E-07 | 158185 | 0.253 | 0.001 | 0.007 | 0.9055 | 474791 |
| rs76550717 | *ARAP1 (CENTD2)* | 11 | 72428172 | A | G | 0.83 | 0.096 | 0.016 | 3.80E-09 | 158185 | 0.816 | -0.011 | 0.006 | 0.0929 | 1600432 |
| rs10830963 | *MTNR1B* | 11 | 92708710 | G | C | 0.266 | 0.077 | 0.015 | 1.70E-07 | 152598 | 0.293 | -0.005 | 0.005 | 0.3854 | 1597551 |
| rs11063018 | *CCND2* | 12 | 4288001 | C | T | 0.19 | 0.088 | 0.017 | 1.30E-07 | 158183 | 0.201 | 0 | 0.006 | 0.965 | 1600265 |
| rs4238013 | *CCND2* | 12 | 4376089 | C | T | 0.2 | 0.099 | 0.017 | 3.60E-09 | 158183 | 0.212 | 0.014 | 0.008 | 0.0724 | 1289047 |
| rs7953190 | *KLHDC5* | 12 | 27962719 | T | C | 0.803 | 0.078 | 0.015 | 4.20E-07 | 158183 | 0.808 | 0.017 | 0.006 | 0.0067 | 1602617 |
| rs2258238 | *HMGA2* | 12 | 66221060 | T | A | 0.1 | 0.11 | 0.02 | 1.60E-07 | 158183 | 0.097 | -0.003 | 0.008 | 0.7191 | 1602612 |
| rs6581998 | *TSPAN8/LGR5* | 12 | 71656723 | C | T | 0.271 | 0.059 | 0.013 | 1.20E-05 | 158183 | 0.259 | 0.001 | 0.006 | 0.9165 | 1605865 |
| rs56348580 | *HNF1A (TCF1)* | 12 | 121432117 | G | C | 0.683 | 0.073 | 0.013 | 2.50E-08 | 158183 | 0.697 | 0.013 | 0.005 | 0.0167 | 1602590 |

| | | | | | | | | | | | | | | | |
|---|---|---|---|---|---|---|---|---|---|---|---|---|---|---|---|
| rs2851437 | *MPHOSPH9* | 12 | 123653592 | A | C | 0.719 | 0.068 | 0.015 | 2.60E-06 | 158183 | 0.744 | -0.005 | 0.006 | 0.3765 | 1600294 |
| rs11616380 | *SPRY2* | 13 | 80705315 | G | T | 0.715 | 0.09 | 0.014 | 3.90E-11 | 158184 | 0.716 | -0.002 | 0.005 | 0.7711 | 1605858 |
| rs10146997 | *NRXN3* | 14 | 79945162 | G | A | 0.212 | 0.068 | 0.015 | 4.60E-06 | 158185 | 0.223 | 0.02 | 0.006 | 0.0006 | 1597551 |
| rs952471 | *HMG20A* | 15 | 77776498 | G | C | 0.687 | 0.082 | 0.013 | 4.00E-10 | 158185 | 0.697 | 0.009 | 0.007 | 0.1634 | 1300374 |
| rs62006309 | *ZFAND6* | 15 | 80411245 | A | G | 0.523 | 0.05 | 0.012 | 4.60E-05 | 158185 | 0.528 | 0.008 | 0.006 | 0.2068 | 1300028 |
| rs12595616 | *PRC1* | 15 | 91563513 | C | T | 0.367 | 0.064 | 0.013 | 5.60E-07 | 152598 | 0.381 | 0.013 | 0.005 | 0.0088 | 1605797 |
| rs1558902* | *FTO* | 16 | 53803574 | A | T | 0.416 | 0.13 | 0.012 | 4.70E-25 | 158182 | 0.414 | 0.051 | 0.005 | 1.55E-25 | 1595482 |
| rs8056814 | *BCAR1* | 16 | 75252327 | G | A | 0.917 | 0.15 | 0.023 | 3.70E-11 | 158182 | 0.916 | 0.021 | 0.009 | 0.0143 | 1607848 |
| rs2925979 | *CMIP* | 16 | 81534790 | T | C | 0.298 | 0.074 | 0.013 | 2.70E-08 | 158182 | 0.304 | 0.003 | 0.005 | 0.5492 | 1599531 |
| rs7224685 | *ZZEF1* | 17 | 4014384 | T | G | 0.304 | 0.068 | 0.013 | 2.00E-07 | 158182 | 0.306 | 0.008 | 0.005 | 0.1342 | 1600601 |
| rs78761021 | *GLP2R* | 17 | 9780387 | G | A | 0.341 | 0.07 | 0.013 | 5.50E-08 | 158182 | 0.336 | 0.015 | 0.005 | 0.0041 | 1602421 |
| rs757209 | *HNF1B (TCF2)* | 17 | 36102833 | G | A | 0.578 | 0.083 | 0.014 | 1.10E-09 | 149849 | 0.547 | -0.002 | 0.005 | 0.6451 | 1595946 |
| rs79349575 | *GIP* | 17 | 46967038 | A | T | 0.506 | 0.065 | 0.013 | 2.60E-07 | 158182 | 0.532 | 0.009 | 0.008 | 0.2459 | 516041 |
| rs7234111 | *LAMA1* | 18 | 7067652 | C | T | 0.364 | 0.063 | 0.013 | 7.70E-07 | 158184 | 0.372 | -0.009 | 0.005 | 0.0728 | 1605845 |
| rs79851087 | *MC4R* | 18 | 58034883 | A | G | 0.972 | 0.17 | 0.041 | 3.20E-05 | 156336 | 0.977 | 0.049 | 0.018 | 0.0049 | 1595703 |
| rs58489806 | *CILP2* | 19 | 19456917 | T | C | 0.091 | 0.082 | 0.021 | 1.00E-04 | 158186 | 0.089 | -0.016 | 0.009 | 0.0574 | 1600481 |
| rs429358 | *APOE* | 19 | 45411941 | T | C | 0.848 | 0.12 | 0.019 | 1.40E-10 | 152543 | 0.841 | 0.015 | 0.007 | 0.023 | 1596844 |
| rs55864746 | *GIPR* | 19 | 46160246 | A | G | 0.309 | 0.065 | 0.014 | 1.80E-06 | 158186 | 0.302 | -0.003 | 0.005 | 0.6092 | 1598865 |
| rs1800961 | *HNF4A* | 20 | 43042364 | T | C | 0.037 | 0.16 | 0.034 | 4.40E-06 | 152595 | 0.038 | -0.009 | 0.013 | 0.4889 | 1585676 |
| rs2023681 | *MTMR3/HORMAD2* | 22 | 30599562 | G | A | 0.888 | 0.12 | 0.021 | 3.90E-09 | 158183 | 0.909 | 0.008 | 0.009 | 0.3296 | 1605896 |

Gene: gene abbreviation of the gene closest to the sentinel SNP; Chr.: Chromosome; Pos.: Position; T2D: Type 2 Diabetes; HF: Heart Failure; EA: Effect Allele; NEA: Non-effect Allele; EAF: Effect Allele Frequency; BETA: Beta coefficient; SE: Standard Error; P: P value; N: sample size.

*Genetic instruments removed in MR-PRESSO.

**Supplementary Table 5.** Summary statistics of two-sample bidirectional MR between T2D and all-cause HF.

| Exposure | Outcome | No. GIVs | IVW-MR | | | MR-Egger | | | |
|---|---|---|---|---|---|---|---|---|---|
| | | | OR | 95% CI | p-value | OR | 95% CI | p-value | Intercept p-value |
| T2D | HF | 82 | **1.07** | **1.04, 1.10** | **$7.02\times10^{-7}$** | 1.01 | 0.94, 1.09 | 0.718 | 0.095 |
| HF | T2D | 59 | **1.6** | **1.36, 1.88** | **$1.55\times10^{-8}$** | **2.05** | **1.20, 3.50** | **0.011** | 0.338 |
| Outliers removed by MR-PRESSO | | | | | | | | | |
| T2D | HF | 77 | **1.06** | **1.03, 1.08** | **$3.33\times10^{-7}$** | 1.02 | 0.95, 1.09 | 0.592 | 0.289 |
| HF | T2D | 57 | **1.42** | **1.26, 1.60** | **$4.85\times10^{-9}$** | **1.5** | **1.02, 2.20** | **0.046** | 0.789 |

GIV: Genetic Instrumental Variable; IVW: inverse variance weighted; MR: Mendelian Randomization; HF: Heart Failure; T2D: Type 2 Diabetes; OR: odds ratio; CI: confidence interval; Bold font: MR association p-values less than 0.05.

**Supplementary Table 6**. The summary of collider bias correction in the GWAS of all-cause heart failure adjusted for type 2 diabetes (genome-wide significant loci $p<5\times10^{-8}$).

| rsID | Gene | Chr. | Pos. (hg19) | EA | NEA | HF GWAS Adjusted for T2D | | | T2D GWAS | | HF GWAS Adjusting for T2D Slope-Hunter Corrected | | Diabetic HF GWAS | | non-Diabetic HF GWAS | |
|---|---|---|---|---|---|---|---|---|---|---|---|---|---|---|---|---|
| | | | | | | **EAF** | **OR (95% CI)** | **P** | **OR (95% CI)** | **P** | **OR (95% CI)** | **P** | **OR (95% CI)** | **P** | **OR (95% CI)** | **P** |
| rs1251863073 | *ZBTB17* | 1 | 16312083 | C | CA | 0.69 | 1.04 (1.03, 1.05) | 2.14E-08 | 1.03 (1.01, 1.04) | 3.21E-05 | 1.05 (1.03, 1.06) | 6.54E-10 | 1.03 (1.01, 1.06) | 0.0019 | 1.04 (1.03, 1.06) | 2.12E-06 |
| rs13376646 | ***C1orf185*** | **1** | **51526232** | **C** | **T** | **0.11** | **1.06 (1.04, 1.08)** | **2.80E-08** | **0.95 (0.93, 0.96)** | **3.99E-08** | **1.05 (1.03, 1.07)** | **5.03E-06** | **1.04 (1.01, 1.08)** | **0.0128** | **1.07 (1.04, 1.1)** | **3.15E-07** |
| rs34517439 | *DNAJB4; GIPC2* | 1 | 78450517 | A | C | 0.11 | 1.06 (1.04, 1.08) | 1.63E-08 | NA | NA | NA | NA | 1.06 (1.03, 1.1) | 0.0003 | 1.06 (1.03, 1.09) | 2.30E-05 |
| rs599839 | *PSRC1* | 1 | 109822166 | A | G | 0.77 | 1.05 (1.03, 1.07) | 1.38E-09 | 1.01 (0.99, 1.02) | 0.4437 | 1.05 (1.03, 1.07) | 1.05E-09 | 1.06 (1.04, 1.09) | 8.11E-07 | 1.04 (1.02, 1.06) | 0.000215 |
| rs10190027 | *STRN* | 2 | 37190726 | C | T | 0.78 | 1.05 (1.03, 1.06) | 9.00E-09 | 1.01 (1, 1.03) | 0.0943 | 1.05 (1.03, 1.07) | 2.88E-09 | 1.04 (1.01, 1.06) | 0.0047 | 1.05 (1.03, 1.08) | 2.59E-07 |
| rs6790914 | *CCDC71* | 3 | 49206318 | G | C | 0.35 | 1.04 (1.02, 1.05) | 3.98E-08 | 1.01 (0.99, 1.02) | 0.2771 | 1.04 (1.03, 1.05) | 2.22E-08 | 1.03 (1.01, 1.05) | 0.0069 | 1.04 (1.03, 1.06) | 7.69E-07 |
| rs2634071 | *NA* | 4 | 111669220 | T | C | 0.21 | 1.07 (1.05, 1.09) | 3.16E-15 | 1.01 (0.99, 1.02) | 0.4114 | 1.07 (1.05, 1.09) | 2.52E-15 | 1.07 (1.04, 1.09) | 1.22E-06 | 1.07 (1.05, 1.09) | 6.81E-10 |
| rs9349379 | *PHACTR1* | 6 | 12903957 | G | A | 0.4 | 1.04 (1.03, 1.05) | 2.67E-09 | 0.99 (0.98, 1) | 0.1125 | 1.04 (1.02, 1.05) | 1.90E-08 | 1.04 (1.02, 1.06) | 2.81E-05 | 1.04 (1.02, 1.05) | 3.08E-05 |
| rs6900627 | *HDGFL1* | 6 | 22570245 | A | G | 0.28 | 1.04 (1.03, 1.06) | 1.50E-09 | 1.01 (1, 1.02) | 0.1819 | 1.05 (1.03, 1.06) | 6.49E-10 | 1.04 (1.02, 1.07) | 0.0001 | 1.04 (1.02, 1.06) | 5.90E-06 |
| rs3176326 | *CDKN1A* | 6 | 36647289 | G | A | 0.8 | 1.07 (1.06, 1.09) | 7.62E-18 | 1.02 (1.01, 1.04) | 0.002 | 1.08 (1.06, 1.1) | 2.81E-19 | 1.05 (1.03, 1.08) | 7.37E-05 | 1.09 (1.07, 1.11) | 4.46E-15 |
| rs10455872 | *LPA* | 6 | 161010118 | G | A | 0.07 | 1.12 (1.09, 1.15) | 2.38E-20 | 1 (0.97, 1.02) | 0.7147 | 1.12 (1.09, 1.15) | 1.31E-19 | 1.11 (1.06, 1.15) | 2.15E-07 | 1.13 (1.1, 1.17) | 1.81E-14 |
| rs73238153 | *FLNC* | 7 | 128481915 | G | A | 0.94 | 1.09 (1.06, 1.12) | 1.81E-09 | 0.96 (0.94, 0.99) | 0.0036 | 1.08 (1.05, 1.11) | 4.20E-08 | 1.08 (1.04, 1.13) | 0.0002 | 1.09 (1.05, 1.13) | 2.24E-06 |
| rs4977575 | *RP11-145E5.5* | 9 | 22124744 | G | C | 0.5 | 1.07 (1.05, 1.08) | 4.42E-23 | 1.02 (1.01, 1.03) | 0.0017 | 1.07 (1.06, 1.08) | 1.12E-24 | 1.06 (1.04, 1.08) | 2.40E-08 | 1.07 (1.05, 1.09) | 4.63E-16 |
| rs2177843 | *SYNPO2L* | 10 | 75409877 | C | T | 0.85 | 1.06 (1.04, 1.08) | 4.80E-10 | 1 (0.98, 1.02) | 0.7979 | 1.06 (1.04, 1.08) | 1.03E-09 | 1.07 (1.04, 1.1) | 4.44E-06 | 1.05 (1.03, 1.08) | 2.15E-05 |
| rs7903146 | ***TCF7L2*** | **10** | **114758349** | **C** | **T** | **0.7** | **1.07 (1.06, 1.09)** | **7.70E-22** | **0.78 (0.77, 0.79)** | **7.65E-319** | **1.03 (1.01, 1.04)** | **1.61E-03** | **1.07 (1.04, 1.09)** | **2.32E-09** | **1.07 (1.05, 1.09)** | **1.17E-13** |
| rs7131546 | *SBF2* | 11 | 10127431 | C | G | 0.12 | 1.06 (1.04, 1.08) | 1.42E-08 | 1.01 (0.99, 1.02) | 0.5434 | 1.06 (1.04, 1.08) | 1.24E-08 | 1.08 (1.04, 1.11) | 1.71E-06 | 1.04 (1.02, 1.07) | 0.000977 |

| | | | | | | | | | | | | | | | | |
|---|---|---|---|---|---|---|---|---|---|---|---|---|---|---|---|---|
| rs35038967 | *BDNF* | 11 | 27703480 | T | A | 0.8 | 1.05 (1.03, 1.07) | 4.80E-09 | 1.01 (1, 1.03) | 0.14854 | 1.05 (1.03, 1.07) | 1.89E-09 | 1.06 (1.03, 1.08) | 1.36E-05 | 1.04 (1.02, 1.06) | 0.000103 |
| rs72805613 | *FTO* | 16 | 53837342 | G | A | 0.41 | 1.04 (1.03, 1.05) | 8.13E-09 | 1.12 (1.11, 1.14) | 1.31E-78 | 1.06 (1.05, 1.07) | 5.24E-17 | 1.04 (1.02, 1.07) | 1.45E-05 | 1.03 (1.01, 1.05) | 0.000258 |
| rs408067 | *SRR* | 17 | 2207236 | C | G | 0.39 | 1.04 (1.03, 1.05) | 8.80E-09 | 1 (0.99, 1.01) | 0.8702 | 1.04 (1.03, 1.05) | 1.59E-08 | 1.02 (1, 1.04) | 0.0563 | 1.05 (1.03, 1.07) | 4.87E-09 |
| rs12150603 | *PGAP3* | 17 | 37834715 | G | A | 0.35 | 1.04 (1.02, 1.05) | 3.54E-08 | 1.03 (1.01, 1.04) | 5.41E-05 | 1.04 (1.03, 1.06) | 1.18E-09 | 1.03 (1.01, 1.05) | 0.0093 | 1.05 (1.03, 1.06) | 4.77E-07 |
| rs61676547 | *BPTF* | 17 | 65892507 | C | G | 0.2 | 1.05 (1.03, 1.06) | 2.90E-08 | 1.06 (1.04, 1.07) | 1.54E-12 | 1.06 (1.04, 1.07) | 5.43E-11 | 1.04 (1.02, 1.07) | 0.0009 | 1.05 (1.03, 1.07) | 9.76E-06 |
| rs62224014 | *LTN1* | 21 | 30347919 | C | T | 0.06 | 1.09 (1.06, 1.12) | 5.73E-10 | 1.01 (0.98, 1.03) | 0.6053 | 1.09 (1.06, 1.12) | 5.72E-10 | 1.07 (1.03, 1.12) | 0.0011 | 1.1 (1.06, 1.14) | 1.07E-07 |

Gene: gene abbreviation of the gene closest to the sentinel SNP; Chr.: Chromosome; Pos.: Position; T2D: Type 2 Diabetes; HF: Heart Failure; EA: Effect Allele; NEA: Non-effect Allele; EAF: Effect Allele Frequency; OR: Odds Ratio; CI: Confidence Interval; P: P value.

**VA Million Veteran Program:**
**Core Acknowledgement for Publications**
**February 2023**

### MVP Program Office

- Sumitra Muralidhar, Ph.D., Program Director
  US Department of Veterans Affairs, 810 Vermont Avenue NW, Washington, DC 20420
- Jennifer Moser, Ph.D., Associate Director, Scientific Programs
  US Department of Veterans Affairs, 810 Vermont Avenue NW, Washington, DC 20420
- Jennifer E. Deen, B.S., Associate Director, Cohort & Public Relations
  US Department of Veterans Affairs, 810 Vermont Avenue NW, Washington, DC 20420

### MVP Executive Committee

- Co-Chair: Philip S. Tsao, Ph.D.
  VA Palo Alto Health Care System, 3801 Miranda Avenue, Palo Alto, CA 94304
- Co-Chair: Sumitra Muralidhar, Ph.D.
  US Department of Veterans Affairs, 810 Vermont Avenue NW, Washington, DC 20420
- J. Michael Gaziano, M.D., M.P.H.
  VA Boston Healthcare System, 150 S. Huntington Avenue, Boston, MA 02130
- Elizabeth Hauser, Ph.D.
  Durham VA Medical Center, 508 Fulton Street, Durham, NC 27705
- Amy Kilbourne, Ph.D., M.P.H.
  VA HSR&D, 2215 Fuller Road, Ann Arbor, MI 48105
- Shiuh-Wen Luoh, M.D., Ph.D.
  VA Portland Health Care System, 3710 SW US Veterans Hospital Rd, Portland, OR 97239
- Michael Matheny, M.D., M.S., M.P.H.
  VA Tennessee Valley Healthcare System, 1310 24$^{th}$ Ave. South, Nashville, TN 37212
- Dave Oslin, M.D.
  Philadelphia VA Medical Center, 3900 Woodland Avenue, Philadelphia, PA 19104

### MVP Co-Principal Investigators

- J. Michael Gaziano, M.D., M.P.H.
  VA Boston Healthcare System, 150 S. Huntington Avenue, Boston, MA 02130
- Philip S. Tsao, Ph.D.
  VA Palo Alto Health Care System, 3801 Miranda Avenue, Palo Alto, CA 94304

### MVP Core Operations

- Lori Churby, B.S., Director, MVP Regulatory Affairs
  VA Palo Alto Health Care System, 3801 Miranda Avenue, Palo Alto, CA 94304
- Stacey B. Whitbourne, Ph.D., Director, MVP Cohort Management
  VA Boston Healthcare System, 150 S. Huntington Avenue, Boston, MA 02130

- Jessica V. Brewer, M.P.H., Director, MVP Recruitment & Enrollment
  VA Boston Healthcare System, 150 S. Huntington Avenue, Boston, MA 02130
- Shahpoor (Alex) Shayan, M.S., Director, MVP Recruitment and Enrollment Informatics
  VA Boston Healthcare System, 150 S. Huntington Avenue, Boston, MA 02130
- Luis E. Selva, Ph.D., Executive Director, MVP Biorepositories
  VA Boston Healthcare System, 150 S. Huntington Avenue, Boston, MA 02130
- Saiju Pyarajan Ph.D., Director, Data and Computational Sciences
  VA Boston Healthcare System, 150 S. Huntington Avenue, Boston, MA 02130
- Kelly Cho, M.P.H, Ph.D., Director, MVP Phenomics Data Core
  VA Boston Healthcare System, 150 S. Huntington Avenue, Boston, MA 02130
- Scott L. DuVall, Ph.D., Director, VA Informatics and Computing Infrastructure (VINCI)
  VA Salt Lake City Health Care System, 500 Foothill Drive, Salt Lake City, UT 84148
- Mary T. Brophy M.D., M.P.H., Director, VA Central Biorepository
  VA Boston Healthcare System, 150 S. Huntington Avenue, Boston, MA 02130
- MVP Coordinating Centers
  - MVP Coordinating Center, Boston - J. Michael Gaziano, M.D., M.P.H.
    VA Boston Healthcare System, 150 S. Huntington Avenue, Boston, MA 02130
  - MVP Coordinating Center, Palo Alto – Philip S. Tsao, Ph.D.
    VA Palo Alto Health Care System, 3801 Miranda Avenue, Palo Alto, CA 94304
  - MVP Information Center, Canandaigua – Brady Stephens, M.S.
    Canandaigua VA Medical Center, 400 Fort Hill Avenue, Canandaigua, NY 14424
  - Cooperative Studies Program Clinical Research Pharmacy Coordinating Center, Albuquerque – Todd Connor, Pharm.D.; Dean P. Argyres, B.S., M.S.
    New Mexico VA Health Care System, 1501 San Pedro Drive SE, Albuquerque, NM 87108

**MVP Publications and Presentations Committee**

- Co-Chair: Tim Assimes, M.D.
  VA Palo Alto Health Care System, 3801 Miranda Avenue, Palo Alto, CA 94304
- Co-Chair: Adriana Hung, M.D.
  VA Tennessee Valley Healthcare System, 1310 24th Ave. South, Nashville, TN 37212
- Co-Chair: Henry Kranzler, M.D.
  Philadelphia VA Medical Center, 3900 Woodland Avenue, Philadelphia, PA 19104

**MVP Local Site Investigators**

- Samuel Aguayo, M.D., Phoenix VA Health Care System
  650 E. Indian School Road, Phoenix, AZ 85012
- Sunil Ahuja, M.D., South Texas Veterans Health Care System
  7400 Merton Minter Boulevard, San Antonio, TX 78229
- Kathrina Alexander, M.D., Veterans Health Care System of the Ozarks
  1100 North College Avenue, Fayetteville, AR 72703
- Xiao M. Androulakis, M.D., Columbia VA Health Care System
  6439 Garners Ferry Road, Columbia, SC 29209
- Prakash Balasubramanian, M.D., William S. Middleton Memorial Veterans Hospital

2500 Overlook Terrace, Madison, WI 53705
- Zuhair Ballas, M.D., Iowa City VA Health Care System
  601 Highway 6 West, Iowa City, IA 52246-2208
- Jean Beckham, Ph.D., Durham VA Medical Center
  508 Fulton Street, Durham, NC 27705
- Sujata Bhushan, M.D., VA North Texas Health Care System
  4500 S. Lancaster Road, Dallas, TX 75216
- Edward Boyko, M.D., VA Puget Sound Health Care System
  1660 S. Columbian Way, Seattle, WA 98108-1597
- David Cohen, M.D., Portland VA Medical Center
  3710 SW U.S. Veterans Hospital Road, Portland, OR 97239
- Louis Dellitalia, M.D., Birmingham VA Medical Center
  700 S. 19th Street, Birmingham AL 35233
- L. Christine Faulk, M.D., Robert J. Dole VA Medical Center
  5500 East Kellogg Drive, Wichita, KS 67218-1607
- Joseph Fayad, M.D., VA Southern Nevada Healthcare System
  6900 North Pecos Road, North Las Vegas, NV 89086
- Daryl Fujii, Ph.D., VA Pacific Islands Health Care System
  459 Patterson Rd, Honolulu, HI 96819
- Saib Gappy, M.D., John D. Dingell VA Medical Center
  4646 John R Street, Detroit, MI 48201
- Frank Gesek, Ph.D., White River Junction VA Medical Center
  163 Veterans Drive, White River Junction, VT 05009
- Jennifer Greco, M.D., Sioux Falls VA Health Care System
  2501 W 22nd Street, Sioux Falls, SD 57105
- Michael Godschalk, M.D., Richmond VA Medical Center
  1201 Broad Rock Blvd., Richmond, VA 23249
- Todd W. Gress, M.D., Ph.D., Hershel "Woody" Williams VA Medical Center
  1540 Spring Valley Drive, Huntington, WV 25704
- Samir Gupta, M.D., M.S.C.S., VA San Diego Healthcare System
  3350 La Jolla Village Drive, San Diego, CA 92161
- Salvador Gutierrez, M.D., Edward Hines, Jr. VA Medical Center
  5000 South 5th Avenue, Hines, IL 60141
- John Harley, M.D., Ph.D., Cincinnati VA Medical Center
  3200 Vine Street, Cincinnati, OH 45220
- Kimberly Hammer, Ph.D., Fargo VA Health Care System
  2101 N. Elm, Fargo, ND 58102
- Mark Hamner, M.D., Ralph H. Johnson VA Medical Center
  109 Bee Street, Mental Health Research, Charleston, SC 29401
- Adriana Hung, M.D., M.P.H., VA Tennessee Valley Healthcare System
  1310 24th Avenue, South Nashville, TN 37212
- Robin Hurley, M.D., W.G. (Bill) Hefner VA Medical Center
  1601 Brenner Ave, Salisbury, NC 28144
- Pran Iruvanti, D.O., Ph.D., Hampton VA Medical Center
  100 Emancipation Drive, Hampton, VA 23667
- Frank Jacono, M.D., VA Northeast Ohio Healthcare System

10701 East Boulevard, Cleveland, OH 44106

- Darshana Jhala, M.D., Philadelphia VA Medical Center
  3900 Woodland Avenue, Philadelphia, PA 19104
- Scott Kinlay, M.B.B.S., Ph.D., VA Boston Healthcare System
  150 S. Huntington Avenue, Boston, MA 02130
- Jon Klein, M.D., Ph.D., Louisville VA Medical Center
  800 Zorn Avenue, Louisville, KY 40206
- Michael Landry, Ph.D., Southeast Louisiana Veterans Health Care System
  2400 Canal Street, New Orleans, LA 70119
- Peter Liang, M.D., M.P.H., VA New York Harbor Healthcare System
  423 East 23rd Street, New York, NY 10010
- Suthat Liangpunsakul, M.D., M.P.H., Richard Roudebush VA Medical Center
  1481 West 10th Street, Indianapolis, IN 46202
- Jack Lichy, M.D., Ph.D., Washington DC VA Medical Center
  50 Irving St, Washington, D. C. 20422
- C. Scott Mahan, M.D., Charles George VA Medical Center
  1100 Tunnel Road, Asheville, NC 28805
- Ronnie Marrache, M.D., VA Maine Healthcare System
  1 VA Center, Augusta, ME 04330
- Stephen Mastorides, M.D., James A. Haley Veterans' Hospital
  13000 Bruce B. Downs Blvd, Tampa, FL 33612
- Elisabeth Mates M.D., Ph.D., VA Sierra Nevada Health Care System
  975 Kirman Avenue, Reno, NV 89502
- Kristin Mattocks, Ph.D., M.P.H., Central Western Massachusetts Healthcare System
  421 North Main Street, Leeds, MA 01053
- Paul Meyer, M.D., Ph.D., Southern Arizona VA Health Care System
  3601 S 6th Avenue, Tucson, AZ 85723
- Jonathan Moorman, M.D., Ph.D., James H. Quillen VA Medical Center
  Corner of Lamont & Veterans Way, Mountain Home, TN 37684
- Timothy Morgan, M.D., VA Long Beach Healthcare System
  5901 East 7th Street Long Beach, CA 90822
- Maureen Murdoch, M.D., M.P.H., Minneapolis VA Health Care System
  One Veterans Drive, Minneapolis, MN 55417
- James Norton, Ph.D., VA Health Care Upstate New York
  113 Holland Avenue, Albany, NY 12208
- Olaoluwa Okusaga, M.D., Michael E. DeBakey VA Medical Center
  2002 Holcombe Blvd, Houston, TX 77030
- Kris Ann Oursler, M.D., Salem VA Medical Center
  1970 Roanoke Blvd, Salem, VA 24153
- Ana Palacio, M.D., M.P.H., Miami VA Health Care System
  1201 NW 16th Street, 11 GRC, Miami FL 33125
- Samuel Poon, M.D., Manchester VA Medical Center
  718 Smyth Road, Manchester, NH 03104
- Emily Potter, Pharm.D., VA Eastern Kansas Health Care System
  4101 S 4th Street Trafficway, Leavenworth, KS 66048
- Michael Rauchman, M.D., St. Louis VA Health Care System

915 North Grand Blvd, St. Louis, MO 63106

- Richard Servatius, Ph.D., Syracuse VA Medical Center
  800 Irving Avenue, Syracuse, NY 13210
- Satish Sharma, M.D., Providence VA Medical Center
  830 Chalkstone Avenue, Providence, RI 02908
- River Smith, Ph.D., Eastern Oklahoma VA Health Care System
  1011 Honor Heights Drive, Muskogee, OK 74401
- Peruvemba Sriram, M.D., N. FL/S. GA Veterans Health System
  1601 SW Archer Road, Gainesville, FL 32608
- Patrick Strollo, Jr., M.D., VA Pittsburgh Health Care System
  University Drive, Pittsburgh, PA 15240
- Neeraj Tandon, M.D., Overton Brooks VA Medical Center
  510 East Stoner Ave, Shreveport, LA 71101
- Philip Tsao, Ph.D., VA Palo Alto Health Care System
  3801 Miranda Avenue, Palo Alto, CA 94304-1290
- Gerardo Villareal, M.D., New Mexico VA Health Care System
  1501 San Pedro Drive, S.E. Albuquerque, NM 87108
- Agnes Wallbom, M.D., M.S., VA Greater Los Angeles Health Care System
  11301 Wilshire Blvd, Los Angeles, CA 90073
- Jessica Walsh, M.D., VA Salt Lake City Health Care System
  500 Foothill Drive, Salt Lake City, UT 84148
- John Wells, Ph.D., Edith Nourse Rogers Memorial Veterans Hospital
  200 Springs Road, Bedford, MA 01730
- Jeffrey Whittle, M.D., M.P.H., Clement J. Zablocki VA Medical Center
  5000 West National Avenue, Milwaukee, WI 53295
- Mary Whooley, M.D., San Francisco VA Health Care System
  4150 Clement Street, San Francisco, CA 94121
- Allison E. Williams, N.D., Ph.D., R.N, Bay Pines VA Healthcare System
  10,000 Bay Pines Blvd Bay Pines, FL 33744
- Peter Wilson, M.D., Atlanta VA Medical Center
  1670 Clairmont Road, Decatur, GA 30033
- Junzhe Xu, M.D., VA Western New York Healthcare System
  3495 Bailey Avenue, Buffalo, NY 14215-1199
- Shing Shing Yeh, Ph.D., M.D., Northport VA Medical Center
  79 Middleville Road, Northport, NY 11768
